# Phenome-wide Mendelian randomization identifying circulating proteins for cardiovascular traits in populations of African ancestry

**DOI:** 10.1101/2025.06.10.25329388

**Authors:** Susannah Selber-Hnatiw, Katerina Trajanoska, Justin Pelletier, Chen-Yang Su, Peyton McClelland, Daniel Taliun, Satoshi Yoshiji, Vincent Mooser, Claude Bhérer, Sirui Zhou

## Abstract

**Background:** Circulating proteins represent robust drug targets with therapeutic potential. Many discoveries have focused on European-ancestry populations, disregarding minuscule yet substantial proteomic differences that may contribute to disease and/or alter drug generalizability in other ancestry groups.

**Methods:** Using two-sample Mendelian randomization and colocalization, we analyzed the effects of 1,556 circulating proteins on 145 cardiometabolic centric outcomes to identify robust protein-phenotype associations in African-ancestry populations and reveal African-ancestry associations with heterogenous effects. We further replicated these findings using the proteomic data available from the UK Biobank Pharma Proteomics Project (UKB-PPP), and tested the effect of protein quantity in association with select phenotypes. Population branch statistics (PBS) were also constructed to examine whether protein-genetic instruments under natural selection could lead to significant protein-outcome associations specific to the African ancestry.

**Results:** We identified 115 robust protein-phenotype associations in African-ancestry populations. Among these, 52 demonstrated heterogenous effects between African- and European-ancestry populations. We further replicated four cross-platform African-ancestry associations in the UKB-PPP and also revealed four significant, direct associations between protein levels and phenotypes. Ultimately, based on our prioritization criteria, we found that CD36, APOC1, GSTA1, and FOLH1, were shown to influence lipids and heart diseases and were uniquely represented in African-ancestry populations. In addition, using PBS, we showed that 47.5% of the 115 significant protein-outcome associations were possibly driven by *cis*-acting-protein quantitative trait loci under natural selection.

**Conclusions:** Multiple lines of evidence were used to interrogate proteomic determinants of cardiometabolic diseases and traits in African-ancestry populations. We highlighted actionable circulating protein targets that could represent potential drug targets for cardiovascular diseases specific to populations with African ancestry.

## Introduction

In the -omics cascade, circulating proteins can be used to proxy biological processes, enabling investigations of the pathophysiological mechanisms of traits and diseases. Over the past decade, multiple lines of evidence have corroborated the utility of circulating proteins in identifying modifiable risk factors, biomarkers, and drug targets ^1–6^. In this context, quantitative trait loci (QTLs) associated with the variation of a phenotype through genetic and/or environmental effects can be used to identify disease associations and support the drug-discovery pipeline ^7^. Leveraging thousands of protein QTLs (pQTLs) uncovered from large genome-wide association studies (GWAS) of the blood proteome, we can identify risk factors for a wide range of diseases ^5,7^ and broaden our understanding of the mechanisms underlying disease influenced by genetic architecture ^8–11^, gene-protein associations ^12–14^, as well as genetic diversity and variation ^14–16^. Recently, there has been an increased number of studies linking the blood proteome to diseases, including Alzheimer’s disease ^17,18^, cardiovascular diseases ^19,20^, and COVID-19 ^4,21–23^. However, most studies have been focused on European-ancestry populations. In contrast, studies of complex diseases more prevalent in African-ancestry populations, such as cardiovascular diseases, diabetes, hypertension, and chronic respiratory infections ^24–27^, that may be explained by ancestry-specific proteomic determinants, are limited ^5,28^. Hence, there is an urgent unmet clinical need to identify the causal and druggable proteomic determinants influencing the health of African-ancestry populations, as well as to devise treatment opportunities for health management and inform on their generalizability.

Mendelian randomization (MR) represents a convenient and powerful tool to assess causality ^29–31^. Similar to the basis of a randomized controlled trial, MR leverages genetic variants that are inherited at random to compare the inter-individual consequences of genetic variants on numerous health outcomes ^32^. Using GWAS summary statistics, investigations of causally associated genetic variants from non-overlapping samples (*i.e.,* two-sample MR) can be performed. To ensure robust causal associations and to avoid bias as well as pleiotropy, three assumptions must be met, namely that genetic variants must be strongly associated with the modifiable risk factor, must only influence the outcome through the modifiable risk factor, and must not be associated with confounders ^30,33^. Circulating proteins represent ideal modifiable risk factors due to their accessibility in the blood and inherent activity in disease ^3,4,34,35^. In particular, *cis*-acting pQTLs (*cis*-pQTLs) can be used as instruments for proteomic MR because they encompass several features that make them an ideal unit for investigating causality. These include their prominence as the most robust genetic marker for proteins ^4,34,36^ and their partial fulfillment of most MR assumptions ^37^, as *cis*-variants are less likely to violate the independence assumption through horizontal pleiotropy ^33^.

Recent studies on disease etiology in diverse populations have revealed an expanded *cis*-genetic architecture of the plasma proteome. Among the *cis*-pQTLs identified and tested for causal associations with diseases, numerous have exhibited clinical trial evidence or drug-repurposing opportunities ^28,38,39^. Moreover, *cis*-pQTLs have been identified in multiple-ancestry populations, with some overlapping between groups and some unique to each population ^5,28,40,41^. The demonstration of this proteogenomic heterogeneity between populations is critical to the drug-development pipeline as the generalizability of drugs is broadly unknown ^42,43^. As such, further investigations are required to properly distinguish the clinical implications of *cis*-pQTLs identified in or between ancestry groups to fully benefit from precision medicine. In this study, we investigated the role of causal proteins associated with multiple traits and diseases, particularly those influencing cardiometabolic outcomes, in African-ancestry populations to highlight unique protein-outcome associations, which may represent candidates for new and robust therapeutic targets.

## Methods

### Plasma proteome GWAS and *cis*-pQTL selection

African *cis*-pQTLs were derived from the ARIC study ^5^ — the largest and most robust proteomic GWAS conducted in African-ancestry ancestry (**Table S1**) — and European *cis*-pQTLs were derived from the Fenland study ^44^ (**Table S2**). In each population, whole-blood proteomic profiling was performed using SomaLogic SomaScans. In the ARIC study, 1,556 independent, sentinel *cis*-pQTLs were identified in *cis*-regions defined as ±500 kb of the protein-encoding gene ^5^, which corresponded to 1,540 proteins and 1,618 SOMAmer reagents (SOMAmers). European *cis*-pQTLs were selected from the Fenland study based on following steps: 1) Published independent *cis*-pQTLs (p<5×10^-8^) that are single nucleotide variants (SNV) ^44^ and 2) For proteins without a *cis*-pQTL with p<5×10^-8^, we selected independent nominal SNV *cis*-pQTLs using clumping (r2<0.001, p<1×10^-5^).

### Outcome GWAS

Resources such as the GWAS catalogue ^45^, the Global Biobank Meta-analysis Initiative (GBMI) ^46^, the COVID-19 host genetics initiative ^47,48^, the Million Veteran Program (MVP) (dbGaP accession phs001672.v3.p1) ^49^, and other databases, as well as literature searches were used to obtain the most contemporary and largest possible GWAS summary statistics for diseases and traits in populations of African ancestry, as well as corresponding phenotypes in Europeans. Upon conducting this search, no criteria nor restriction on phenotypes was considered as there are limited numbers of GWAS summary statistics available in diverse populations. GWAS summary statistic files containing minimally essential variables (*e.g.*, rsID or SNP, effect, other, or reference alleles, beta or odds ratio, and p-value) were retained and otherwise discarded if it was not possible to compute and/or transform variables manually.

A total of 145 phenotypes were gathered and analyzed in this study, reflecting African-specific GWAS summary statistics that included minimally essential variables (**Table S3**, **S1 Appendix**). GWAS summary statistics from the following phenotypes were selected, including body mass index (BMI) ^50^, height ^51^, lipid traits (high-density lipoprotein (HDL)^52,53^, low-density lipoprotein (LDL) ^45^, total cholesterol [95], non-HDL cholesterol ^52^ triglyceride levels ^45^), asthma ^54^, primary open-angle glaucoma (POAG) ^55^, smoking status (smoking initiation and cessation) ^56^, stroke (cardioembolic stroke, ischemic stroke, large artery stroke, small vesicle stroke, stroke) ^57^, type 2 diabetes ^58^, chronic obstructive pulmonary disorder (COPD) ^46^, gout ^46^, heart failure (HF) ^46^, idiopathic pulmonary fibrosis (IPF) ^46^, venous thromboembolism (VTE) ^46^, coronavirus disease (COVID-19) status (very severe confirmed COVID-19, hospitalized COVID-19, COVID-19) ^47,48^ post-traumatic stress disorder (PTSD) ^59^, diabetic retinopathy ^60^, as well as 114 cardiovascular related phenotypes from the MVP (**Table S3**, **S1 Appendix**) (dbGaP accession phs001672.v3.p1) ^49^. Moreover, lipid traits were obtained for two African-ancestry populations including Sub-Saharan Africans (POP1), and African Americans, Afro-Caribbeans, African unspecified (POP2) to investigate potential population differences (**S1 Appendix**). A detailed description of each African GWAS summary statistic and phenotype is presented in **Table S3** and **S1 Appendix**. European GWAS summary statistics for the equivalent phenotypes were gathered using the same search methods and criteria. A detailed description of each European GWAS summary statistic and phenotype is presented in **Table S4**).

### Preparation of GWAS summary statistics for Mendelian randomization

As several GWAS summary statistic files were available in GRCh37/hg19 format, we performed liftover of the GRCh38/hg38 files to GRCh37/hg19 for ease of analysis, using the UCSC Genome Browser GRCh38/hg38 or GRCh37/hg19 chain files ^61^. Additionally, variables were computed or transformed when missing from GWAS summary statistic files (*e.g.*, SNP IDs (*i.e.*, rsID), effect allele frequency (EAF), effect size, standard error (SE)). When absent, SNPs were mapped based on chromosome number and position, and EAF was calculated with PLINK v1.9 ^62^ using the 1000 Genomes Project (1KGP) YRI reference panels ^63^.

### Two-sample Mendelian randomization

We used MR to estimate the causal effect of modifiable risk factors (*i.e*., plasma proteins) on disease or traits (*i.e*., outcomes) using instrumental variables (*i.e*., *cis*-pQTLs). A harmonization step was first performed using the R package *TwoSampleMR* v0.5.8 ^64^ to ensure the proper allelic alignment and direction between the SNPs on the exposure and outcome. Ambiguous and unambiguous SNPs, as well as palindromic SNPs, were identified, and variants characterized by strand issues were automatically updated (*e.g.*, flipping effect alleles for the reference when coded on the alternative strand) or removed (*e.g.*, alleles in the exposure and outcome that did not correspond to the same SNP, or were palindromic SNPs with a minor allele frequency (MAF) >0.42).

Using *cis*-pQTLs as instrumental variables, *TwoSampleMR* ^64^ was used to evaluate the causal role of proteins in African-ancestry individuals or in European-ancestry individuals for the selected outcomes. For SNPs that failed the harmonization step due to Linkage disequilibrium (LD), SNP proxies were identified using the tool *Snappy* v1.0 ^65^. Specifically, SNPs were matched by chromosome number and position to those in the 1000 genomes project (1KGP) Yoruba in Ibadan, Nigeria (YRI) reference panel (LD r^2^ > 0.8) for African-ancestry individuals or the EUR reference panel (LD r^2^ > 0.8) for European-ancestry individuals ^63^.

Upon performing two-sample MR, different estimators were used to conduct analyses depending on the number of instrumental variables influencing outcome associations, including the Wald ratio for a single instrumental variable, and the inverse variance-weighted method or MR-Egger method for multiple independent instrumental variables that can concurrently influence outcome associations.

## Colocalization

For significant proteins associated with any of the 145 outcomes in African-ancestry populations that survived FDR corrections (FDR pval<0.05), colocalization was conducted to limit confounding potentially arising from LD. That is, using Bayesian statistical testing, colocalization was used to test whether each pair of independent exposure-outcome associations were shared by the same causal variant, demonstrating correlation. Coloc (v5.2.1) ^66^ was used to assess colocalization, in which correlations were assessed based on the posterior probability for hypothesis four (PP.H4>0.7), defined as when two association signals are likely shared by the same causal variant.

### Replication of proteins in European populations

MR was repeated in Europeans using the same methods as above to test the multi-ancestry effect of proteins on corresponding phenotypes. In doing so, significant African (FDR pval<0.05, PP.H4>0.7) and equivalent European protein-phenotype associations (*i.e.*, protein IDs and outcomes) were compared to distinguish and focus on heterogenous associations between the two populations. To discern heterogeneity — the effect of African proteins on outcomes estimated by MR between African- and European-ancestry populations — the meta-analysis tool METAL ^67^ was used. Two analysis schemes were selected, including the SE analysis scheme and the random effects analysis scheme. First, the SE analysis scheme was selected to combine effect size estimates and SEs. In doing so, METAL was used to compute Chi^2^ and p-values to understand whether there were statistically significant differences between the effect of a MR significant protein between ancestry groups, as well as I^2^ to extrapolate the percentage of variability resulting from effect estimates ^67^. Chi^2^ was calculated based on the weighted sum of squared differences between individual study effects and the pooled effect across studies, p-values were calculated based on the weighted sum from each individual statistic, and I^2^ was calculated using Chi^2^ statistics and degrees of freedom from each individual statistic ^67^. Heterogeneity was observed in the presence of high Chi^2^ values and low p-values, and large estimates of I^2^ (I^2^>75%). Second, considering there is variability between studies (*i.e*., conducted under different conditions with dissimilar participants), a random effects model, METAL-Random v0.1.0 ^68^, was used to reconcile between-study variability. METAL-Random ^68^ uses the DerSimonian-Laird random effects model to calculate Tau^2^ by meta-analyzing the intervention effects and SE from each study ^69^. In this context, heterogeneity with random effects was observed in the presence of the highest Tau^2^ values.

### Protein altering variants, expression and splicing QTLs, and pleiotropy

Protein altering variants (PAVs) potentially influencing aptamer or antibody binding during protein level quantification were examined using HaploReg v4.2 ^70^. If a *cis*-pQTL is a missense, nonsense, or splicing variant, or in LD (R^2^≧0.6) with a missense, nonsense, or splicing variant, it was flagged as a potential PAV. An African 1KGP Phase 1 population was used for LD calculations. GTex was used to identify single-tissue expression QTLs (eQTLs) and splicing QTLs (sQTLs) (see URLs). Phenoscanner and HaploReg v4.2 were used to interpret potential pleiotropic effects ^71^. Inclusion criteria for the final prioritization of protein-outcome associations was based on generalized criteria thought to encompass robust modifiable risk factors and drug targets, including a significant p-value, potential or existing drug targets, described OMIM gene-phenotype relationships ^72^ and/or well-described in the literature. Moreover, *cis*-pQTLs that are PAVs were also included to highlight mutations implicated in disease.

### Replication in the UKB-PPP

Phenotypic and proteomic data from the UK Biobank (UKB) and UK Biobank Pharma Proteomics Project (UKB-PPP) were accessed under project 73958. The UKB is a large-scale biobank containing a total of >500,000 genomes with proteomics examined in >54,000 participants belonging to the UKB-PPP ^7^.

For prioritized proteins and outcomes in African populations, we repeated MR using *cis*-pQTLs identified from the UKB-PPP African ancestry cohort ^7^. *cis*-pQTLs were matched based on corresponding proteins and MR was repeated in the same manner as described above. Using individual-level data from the UKB-PPP, the quantity of protein directly associated with each *cis*-gene was identified. African-ancestry individuals were identified in the UKB-PPP based on Data-Field 21000, which includes Black or Black British, Caribbean, or African individuals, and any other individuals from Black backgrounds. Disease and trait profiles corresponding to the 145 outcome phenotypes were then extracted, including blood biochemistry (Category 17518) for the relevant lipid traits (*e.g.*, total cholesterol, Data-Field 30690; HDL, Data-Field 30760; LDL Data-Field 30780, triglycerides Data-Field 30870), and diseases based on self-reported and/or ICD10 codes (Data-Field 12144).

The R package *glm2* v1.2.1 was used to construct linear regressions ^73^, utilizing the Gaussian function for continuous traits and the binary function for binary traits. Moreover, normalization strategies were applied to datasets, in addition to removing zeroes at baseline and removing standard deviations >5. To improve regression models for continuous phenotypes, covariates were added including sex, age, and smoking status.

### Population branch statistics (PBS)

We calculated the PBS for each *cis*-pQTL from the African populations to infer if any *cis*-pQTLs are operating under natural selection. The PBS represents a fixation metric used to distinguish demographic history by comparing the level of natural selection (*i.e.*, differentiation) observed between individuals of one population to two reference populations based on the allele frequency. In doing so, PBS values were computed on all available *cis*-pQTLs from the ARIC discovery analysis (n=1,540) to understand the level of sequence differentiation between African, European, and East Asian reference populations. PBS values were computed using the 30⨉ whole genome sequencing data from the Human Genome Diversity Project (HGDP) ^63^, in which genetic variation (*i.e.*, changes in allele frequency) was measured for each subgroup and compared to the total level of genetic variation between each population. Integrated haplotype scores were computed using the *allel.pbs* function in Python v3.11.4 using *scikit-allel v1.3.8* ^74^. Absolute value of PBS scores was used in identifying putative natural selection ^75–77^. For any *cis*-pQTLs with a MAF=0 in any of the three populations, one allele is added for calculating PBS.

## Results

### Protein MR results in African-ancestry populations

The study design is displayed in **Figure 1** and a detailed description of the exposure and outcome populations are provided in the **Methods**. Two-sample MR was performed using 1,540 *cis*-pQTLs (located ±500 kb from the transcription start-site) representing 1,556 circulating proteins — originally identified in a previous study (ARIC) ^5^ of 1,871 African-ancestry participants (**Table S1**) — on 145 outcome GWASs (**Table S3**). For *cis*-pQTLs that were not present in the outcome GWASs, a total of 921 unique proxy *cis*-pQTLs (representing 905 proteins across the 145 outcomes) were identified based on LD scores (R^2^>0.8) and genomic coordinates using the 1000 genomes project (1KGP) Yoruba in Ibadan, Nigeria (YRI) reference panel (**Table S5**) ^63^. In total, 133 significant (FDR p<0.05) protein-outcome associations were identified, representing 44 proteins (**Table S6**). The majority of these significant associations related to cardiometabolic traits; specifically, 70 signals were associated with metabolic outcomes, 44 with cardiovascular outcomes, 13 with vascular outcomes, five with anthropometrics, and one with a pulmonary outcome.

**Figure 1:**
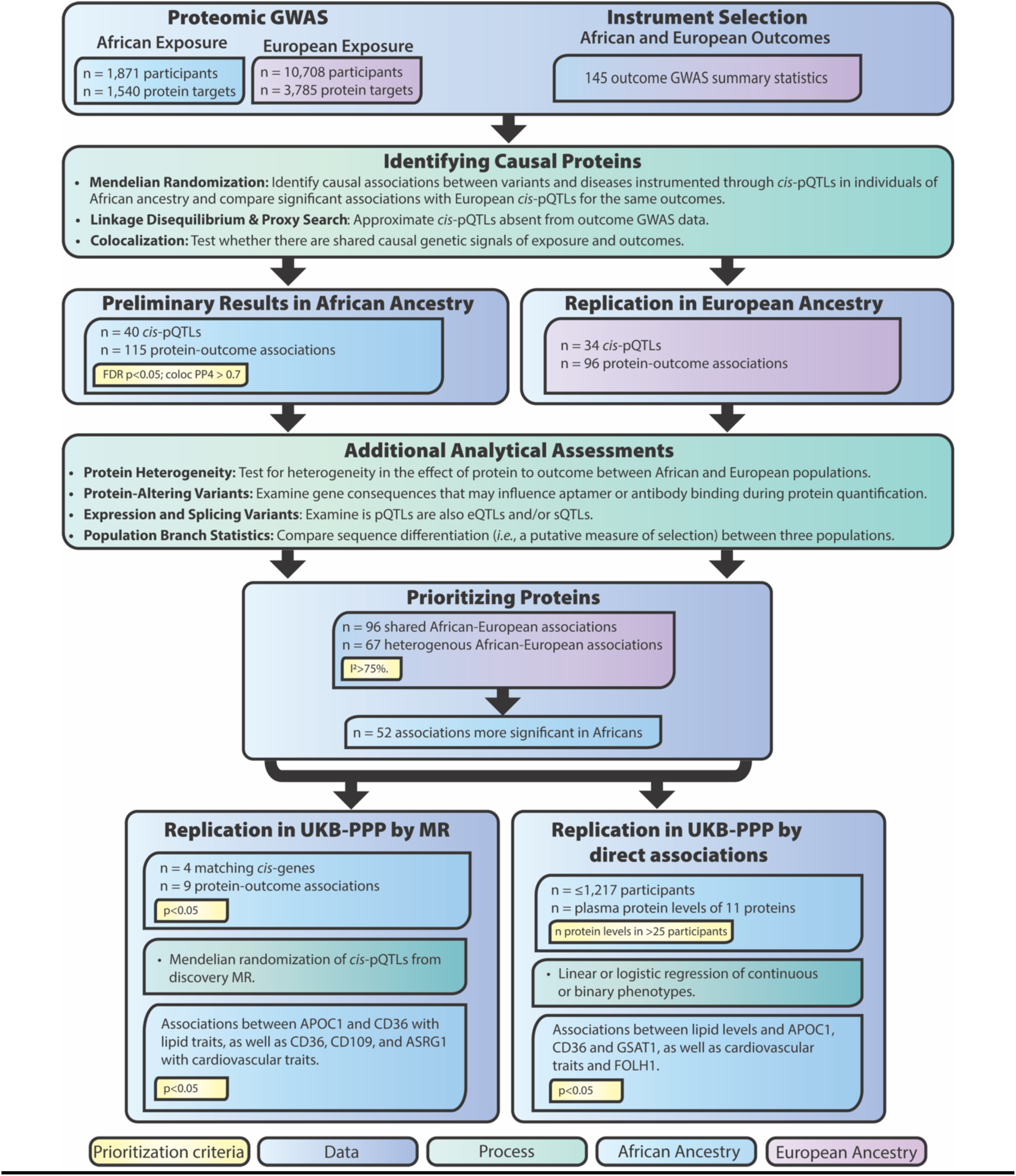
Study overview and workflow displaying prioritization criteria and the successive ranking of causal proteins in Africans based on replication in other populations. pQTL= protein quantitative trait loci; eQTL= expression quantitative trait loci; sQTL= splicing quantitative trait loci; LD= Linkage Disequilibrium; UKB-PPP= UK Biobank Pharma Proteomics Project.

### Colocalization

Colocalization was then conducted using Coloc ^66^ on the 133 significant protein-outcome pairs to identify shared robust causal signals between proteins and outcomes. We considered genetic signals to be colocalized if the posterior probability for a shared causal signal (*i.e*., PP.H4) was more than 0.7 (PP.H4>0.7). In total, genetic signals of 40 proteins (115 protein-outcome associations) were found to colocalize with at least one outcome (**Table S7**).

### Replication in European cohorts

To further prioritize the robust set of causal proteins and identify those which are significantly and uniquely present in African ancestry, we repeated analyses in European-ancestry cohorts, focusing on the 40 proteins (115 protein-outcome associations) prioritized from the previous analysis (**Tables S2 and S8, Figure 1**).

Prioritized protein-outcome associations in the African studies were identified by matching protein SOMAmer IDs (SeqIDs) and outcome associations with the lowest p-value per MR method, including the Wald ratio, inverse-variance-weighted, weighted median, simple mode, and MR-Egger in the European studies (**Tables S2 and S9**). A total of 96 SeqID-outcome associations significant in the African studies were found in the European studies, while 18 SeqID-outcome associations significant in African studies were absent in European studies and not considered in further analyses. Two-sample MR was then performed using 96 *cis*-pQTLs (**Table S9**), representing 34 proteins — originally identified in a previous study of 10,708 European-ancestry participants ^44^ — on the same 145 outcome GWASs (**Tables S3 and S4**). Proxies were identified for *cis*-pQTLs not present in the outcome GWASs, which included 3 *cis*-pQTLs representing 3 proteins based on LD scores (R^2^>0.8) and genomic coordinates in the 1KGP European population samples (**Table S9**) ^63^.

For the 96 shared SeqID-outcome associations in the African and European studies, protein-outcome heterogeneity was tested using METAL ^67^ and METAL-Random v0.1.0^68^. In total, 67 out of 96 (69.79%) shared associations demonstrated high heterogeneity (I^2^>75%) with additive random effects (τ^2^) ranging from 1.20×10^-4^ to 3.86×10^-1^, depicting varying levels of between-study variation (**Table S9**). Among these, a subset of 52 associations were more significant in Africans than in Europeans and were therefore prioritized for further study (**Table S10, Figure 2**). Particularly, 39 of the 52 associations (representing 17 proteins), were non-significant in the European populations. Among prioritized associations, 50 out of 52 were enriched for traits associated with cardiovascular disorders.

**Figure 2:**
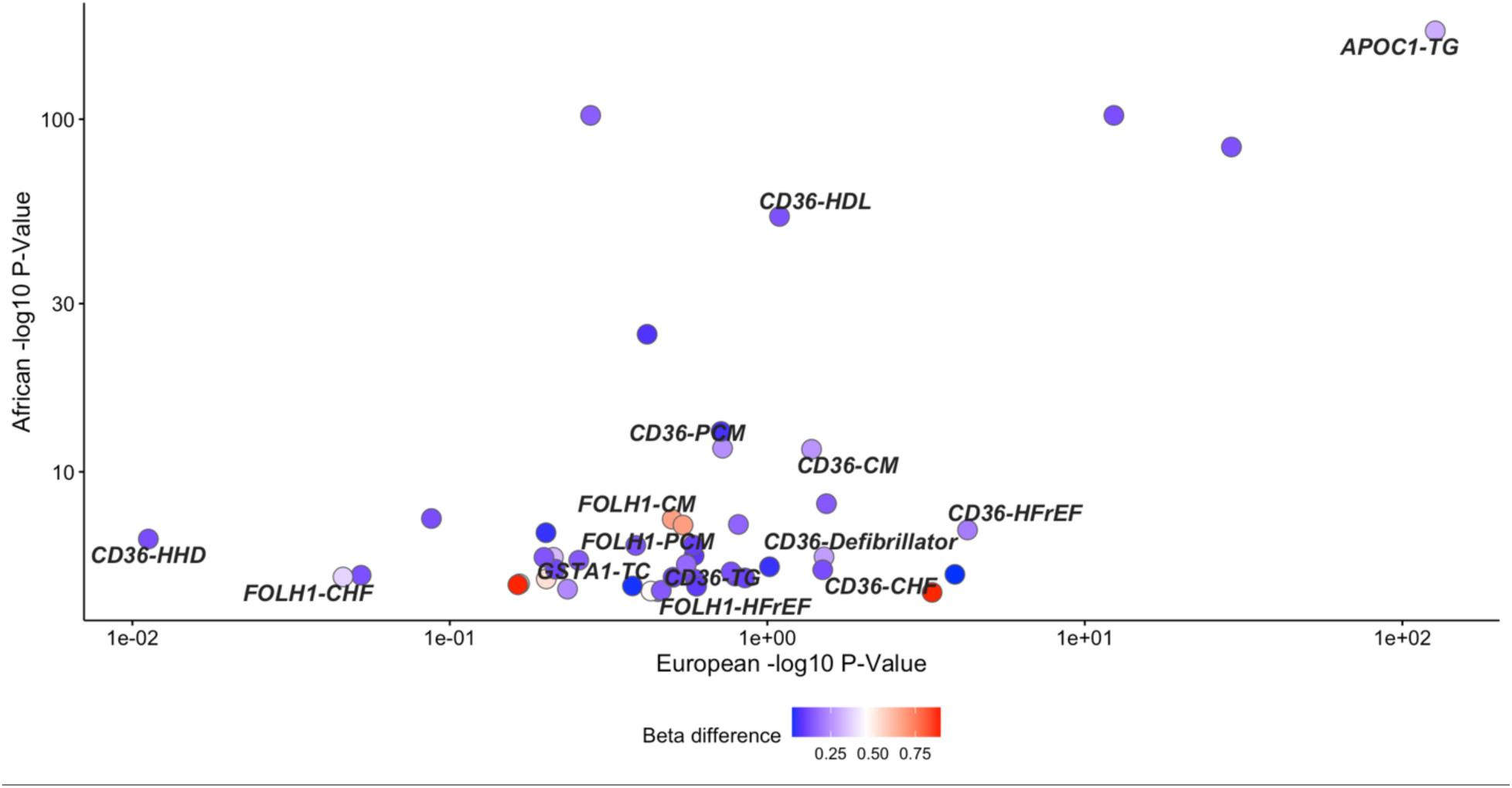
Plot showing differences in absolute effect sizes between African and European populations. Demonstrated in text boxes are differences in absolute effect size for top prioritized protein-outcome associations (**Table 2 and 3**). **Notes**: TG = Triglyceride levels; TC = Total cholesterol; Defibrillator = Cardiac defibrillator in situ; CM = Cardiomyopathy; CHF = Congestive heart failure (non-hypertensive); HDL = High-density lipoproteins; HFrEF = Heart failure with reduced ejection fraction [Systolic or combined heart failure]; HHD = Hypertensive heart disease; PCM = Primary/intrinsic cardiomyopathies; CHF = Congestive heart failure (non-hypertensive).

### Converging lines of evidence on potential protein biomarkers in African-ancestry populations

The 52 significant associations were further scrutinized, quantifying them based on the statistical analyses (FDR p<0.05, PP.H4>0.7 in African ancestry, and I^2^>75% between African and European studies, **Table S10**) as well as qualitative evidence such as potential or existing drug targets, descriptions of known OMIM gene-phenotype relationships ^72^, or other evidence in the literature (**Figure 1**). Several proteins were significant for multiple outcomes, particularly for lipid traits (**Table 1**), and some exhibited possible pleiotropy (**Table S11**) or possible pleiotropy based on variants in LD with the *cis*-pQTL (**Table S12**). Moreover, 20 *cis*-pQTLs were also eQTLs (**Table S13**), and 10 *cis*-pQTLs were also sQTLs (**Table S14**). Corresponding results in the European population can be found in **S2 Appendix**.

**Table 1.**
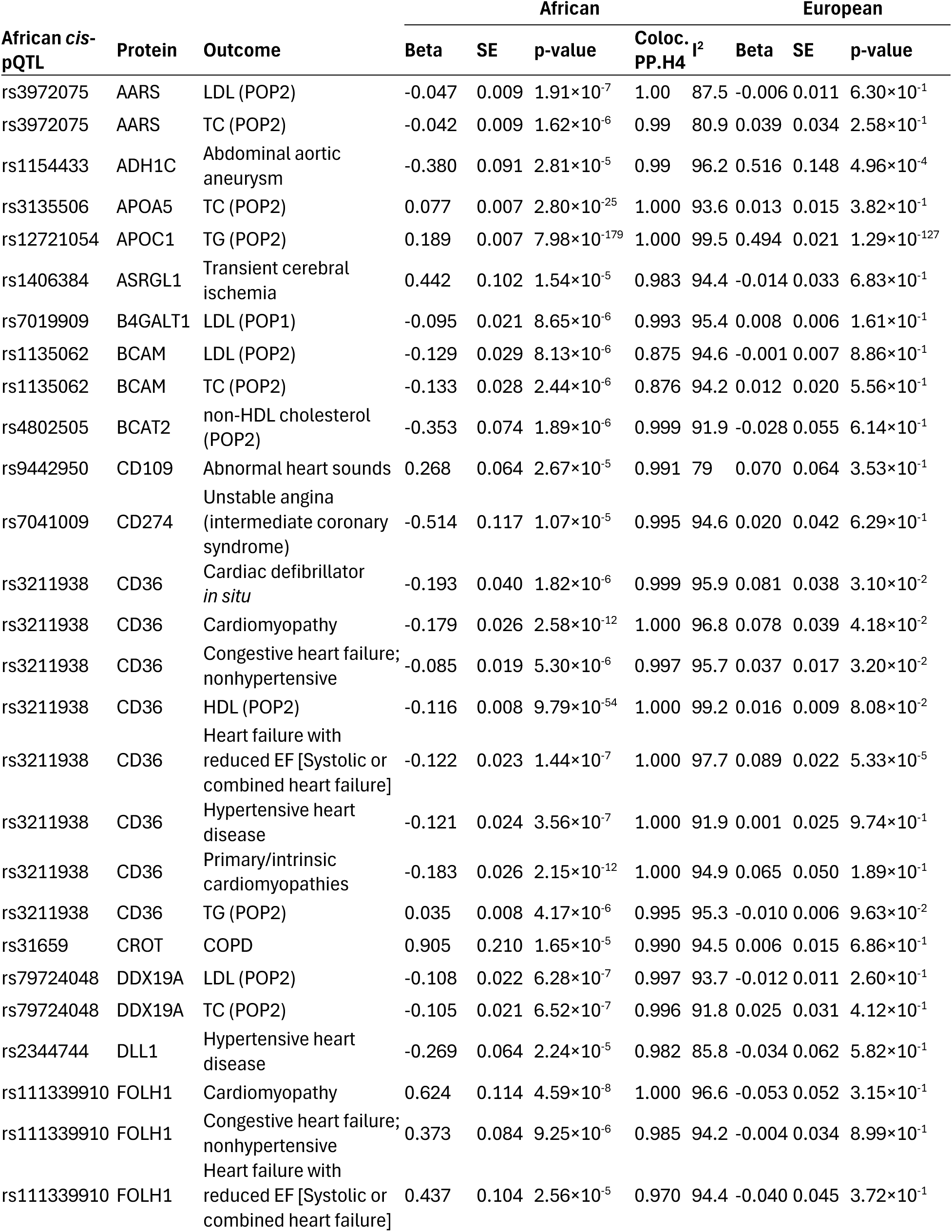
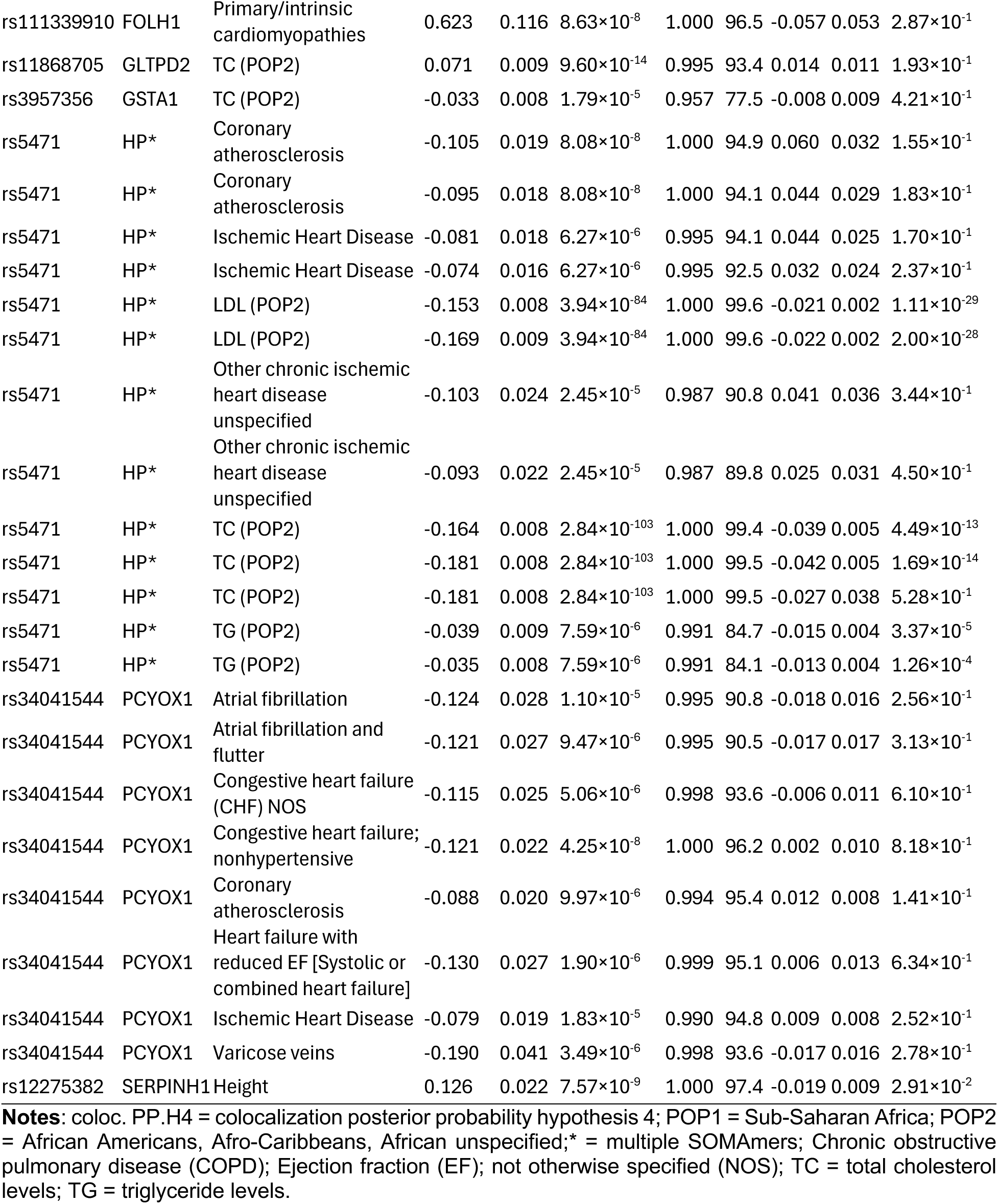
Prioritized protein-outcome associations in African ancestry populations by MR.

**Table 2.**
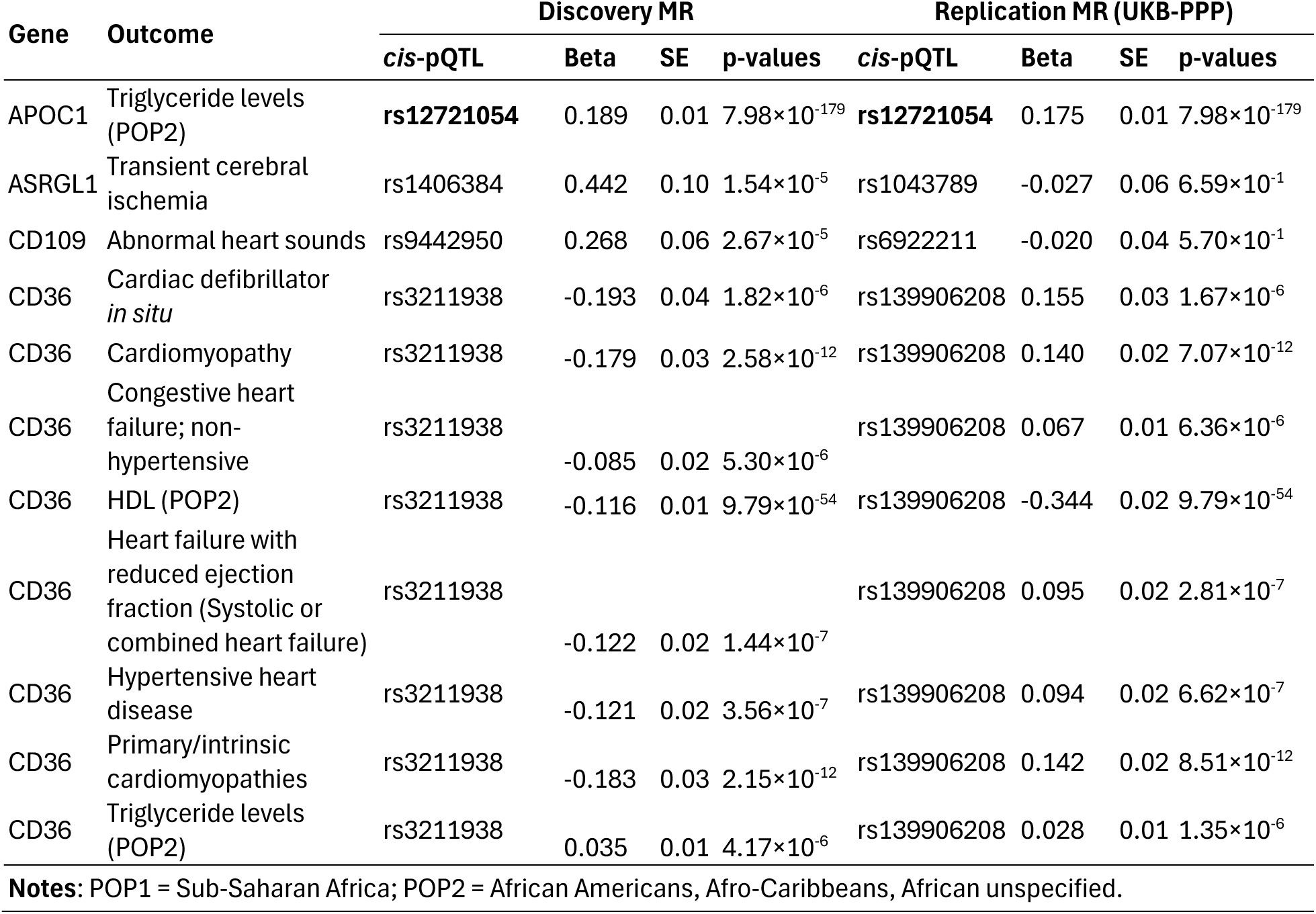
Cross-platform MR results of prioritized protein-outcome associations in African ancestry populations.

| Gene | Outcome | Discovery MR |  |  |  | Replication MR (UKB-PPP) |  |  |  |
| --- | --- | --- | --- | --- | --- | --- | --- | --- | --- |
|  |  | *cis*-pQTL | Beta | SE | p-values | *cis*-pQTL | Beta | SE | p-values |
| APOC1 | Triglyceride levels (POP2) | **rs12721054** | 0.189 | 0.01 | $7.98 \times 10^{-179}$ | **rs12721054** | 0.175 | 0.01 | $7.98 \times 10^{-179}$ |
| ASRGL1 | Transient cerebral ischemia | rs1406384 | 0.442 | 0.10 | $1.54 \times 10^{-5}$ | rs1043789 | -0.027 | 0.06 | $6.59 \times 10^{-1}$ |
| CD109 | Abnormal heart sounds | rs9442950 | 0.268 | 0.06 | $2.67 \times 10^{-5}$ | rs6922211 | -0.020 | 0.04 | $5.70 \times 10^{-1}$ |
| CD36 | Cardiac defibrillator *in situ* | rs3211938 | -0.193 | 0.04 | $1.82 \times 10^{-6}$ | rs139906208 | 0.155 | 0.03 | $1.67 \times 10^{-6}$ |
| CD36 | Cardiomyopathy | rs3211938 | -0.179 | 0.03 | $2.58 \times 10^{-12}$ | rs139906208 | 0.140 | 0.02 | $7.07 \times 10^{-12}$ |
| CD36 | Congestive heart failure; non-hypertensive | rs3211938 | -0.085 | 0.02 | $5.30 \times 10^{-6}$ | rs139906208 | 0.067 | 0.01 | $6.36 \times 10^{-6}$ |
| CD36 | HDL (POP2) | rs3211938 | -0.116 | 0.01 | $9.79 \times 10^{-54}$ | rs139906208 | -0.344 | 0.02 | $9.79 \times 10^{-54}$ |
| CD36 | Heart failure with reduced ejection fraction (Systolic or combined heart failure) | rs3211938 | -0.122 | 0.02 | $1.44 \times 10^{-7}$ | rs139906208 | 0.095 | 0.02 | $2.81 \times 10^{-7}$ |
| CD36 | Hypertensive heart disease | rs3211938 | -0.121 | 0.02 | $3.56 \times 10^{-7}$ | rs139906208 | 0.094 | 0.02 | $6.62 \times 10^{-7}$ |
| CD36 | Primary/intrinsic cardiomyopathies | rs3211938 | -0.183 | 0.03 | $2.15 \times 10^{-12}$ | rs139906208 | 0.142 | 0.02 | $8.51 \times 10^{-12}$ |
| CD36 | Triglyceride levels (POP2) | rs3211938 | 0.035 | 0.01 | $4.17 \times 10^{-6}$ | rs139906208 | 0.028 | 0.01 | $1.35 \times 10^{-6}$ |

**Notes:** POP1 = Sub-Saharan Africa; POP2 = African Americans, Afro-Caribbeans, African unspecified.

### Cross-platform MR replication

To test the robustness of our findings, we repeated the MR analysis of the 52 prioritized protein-outcome associations (using proteomic GWAS from ARIC, Somalogic platform) using genetic instruments derived from the proteomic GWAS from the UKB-PPP (Olink platform) ^44^. The UKB-PPP comprises a large proteomic resource useful for interrogating disease etiology and validating therapeutic targets. The UKB-PPP was built using several Olink protein panels including cardiometabolic, inflammation, neurology, and oncology ^7^.

A total of 4 proteins with *cis*-pQTL from the UKB-PPP African-ancestry GWAS were found among the 52 prioritized associations ^7^, where 3 out of 4 proteins had different *cis*-pQTLs between the ARIC proteomic GWAS and UKB-PPP GWAS (**Table S15**). Two-sample MR was then performed using the 4 proteins from the UKB-PPP GWAS as exposures on matching outcomes from the discovery analysis, which yielded a total of 11 protein-outcome associations (**Table S16**). Overall, 9 of 11 associations were replicated using the UKB-PPP African proteomic GWAS (p<0.05) (**Table 2, Table S16**), including APOC1,CD109, ASRGL1, and multiple significant associations for CD36 (**Table 2, Figure 2, Figure S1**). Notably, one *cis*-pQTL was identical between proteomic GWASs using measurements from SomaLogic and Olink platforms, yielding the same result (in bold, **Table 2**).

### Associations between plasma protein levels and phenotypic outcomes in African-ancestry populations

Using the individual measurements from the UKB, we examined associations between measured plasma protein levels and the phenotypes of interest in 52 prioritized associations from MR. To identify phenotypes in the UKB, GWAS outcome-phenotype definitions were reviewed to elucidate the best matching ICD10 codes (if not provided), or direct traits were used (*e.g.*, lipid levels measured from the serum) (**Table S17**). Nine of the 20 proteins in the prioritized associations were measured in the UKB-PPP (**Table S18**).

The UKB contained 8,058 African-ancestry participants (see definitions in **Methods**), with proteomic profiling in 1,217 African-ancestry participants in the UKB-PPP ^7^. Linear regression was performed on relevant continuous traits using individual protein quantification data from the UKB-PPP, including 1,090 participants with reported APOC1 triglyceride proteins, 1,175 participants with reported B4GALT1 proteins for LDL levels, 1,175 participants with reported BCAM proteins for both LDL and total cholesterol levels, 1,058 participants with reported BCAT2 proteins for LDL levels, 1,166 participants with reported GSTA1 proteins for total cholesterol levels, 1,073 participants with reported SERPINH1 proteins for height measurements, and 1,062 participants with reported CD36 proteins for both HDL and triglyceride levels (**Table 3**).

**Table 3.** Direct associations of prioritized plasma protein levels and outcomes in the UKB-PPP.

|  | Protein | Outcome (ICD10) | Participants (n) | Coefficients (CI) | P-value |
| --- | --- | --- | --- | --- | --- |
| Continuous | APOC1 | Triglycerides | 1090 | 0.166 (0.102, 0.229) | 3.51×10<sup>-7</sup> |
|  | B4GALT1 | LDL | 1175 | -0.009 (-0.066, 0.048) | 7.52×10<sup>-1</sup> |
|  | BCAM | LDL | 1175 | -0.024 (-0.08, 0.033) | 4.13×10<sup>-1</sup> |
|  | BCAT2 | LDL | 1058 | 0.003 (-0.058, 0.064) | 9.24×10<sup>-1</sup> |
|  | CD36 | Triglycerides | 1062 | 0.198 (0.134, 0.263) | 2.43×10<sup>-9</sup> |
|  | CD36 | HDL | 1062 | -0.062 (-0.126, 0.002) | 5.94×10<sup>-2</sup> |
|  | GSTA1 | Cholesterol | 1166 | 0.098 (0.042, 0.154) | 6.29×10<sup>-4</sup> |
|  | SERPINH1 | Height | 1073 | 0.027 (-0.057, 0.111) | 5.27×10<sup>-1</sup> |
| Binary | CD274 | Unstable angina<br>(intermediate coronary syndrome) | 55 cases,<br>1106 controls | 0.621 (-0.05, 1.30) | 7.12×10<sup>-2</sup> |
|  | CD36 | Congestive heart failure; non-hypertensive | 49 cases,<br>1011 controls | -0.658 (-1.50, 0.183) | 1.25×10<sup>-1</sup> |
|  | FOLH1 | Congestive heart failure; non-hypertensive | 49 cases,<br>1011 controls | 1.798 (1.12, 2.48) | 2.20×10<sup>-7</sup> |

Logistic regression was then performed on relevant binary traits using individual protein quantification data from the UKB-PPP, including 55 participants with reported CD274 proteins for unstable angina (intermediate coronary syndrome), 49 participants with reported CD36 proteins for both congestive heart failure (non-hypertensive) and heart failure with reduced EF (Systolic or combined heart failure), as well as 49 participants with reported FOLH1 proteins for both congestive heart failure (non-hypertensive) and heart failure with reduced EF (Systolic or combined heart failure). Traits with less than 25 participants of African ancestry in the prioritized associations were not tested due to insufficient power.

We identified significant associations between triglyceride levels and APOC1 (p=3.51×10^-7^) as well as CD36 (p=2.43×10^-9^), total cholesterol levels and GSTA1 (p=6.29×10^-4^), congestive heart failure (non-hypertensive) and FOLH1 (p=2.20×10^-7^) (**Figure 3, Figure S2**), and suggestive associations between HDL levels and CD36 (p=5.94×10^-2^), as well as unstable angina (intermediate coronary syndrome) and CD274 (p=7.12×10^-2^) (**Table S18, Figure S2**). No significant associations were observed between LDL levels and B4GALT1, BCAM, or BCAT2 (p=0.793, p=0.568, p=309, respectively) (**Figure S2**), height and SERPINH1 (p=0.153), nor congestive heart failure (non-hypertensive) and CD36 (p=0.125) (**Figure S3**).

**Figure 3:**
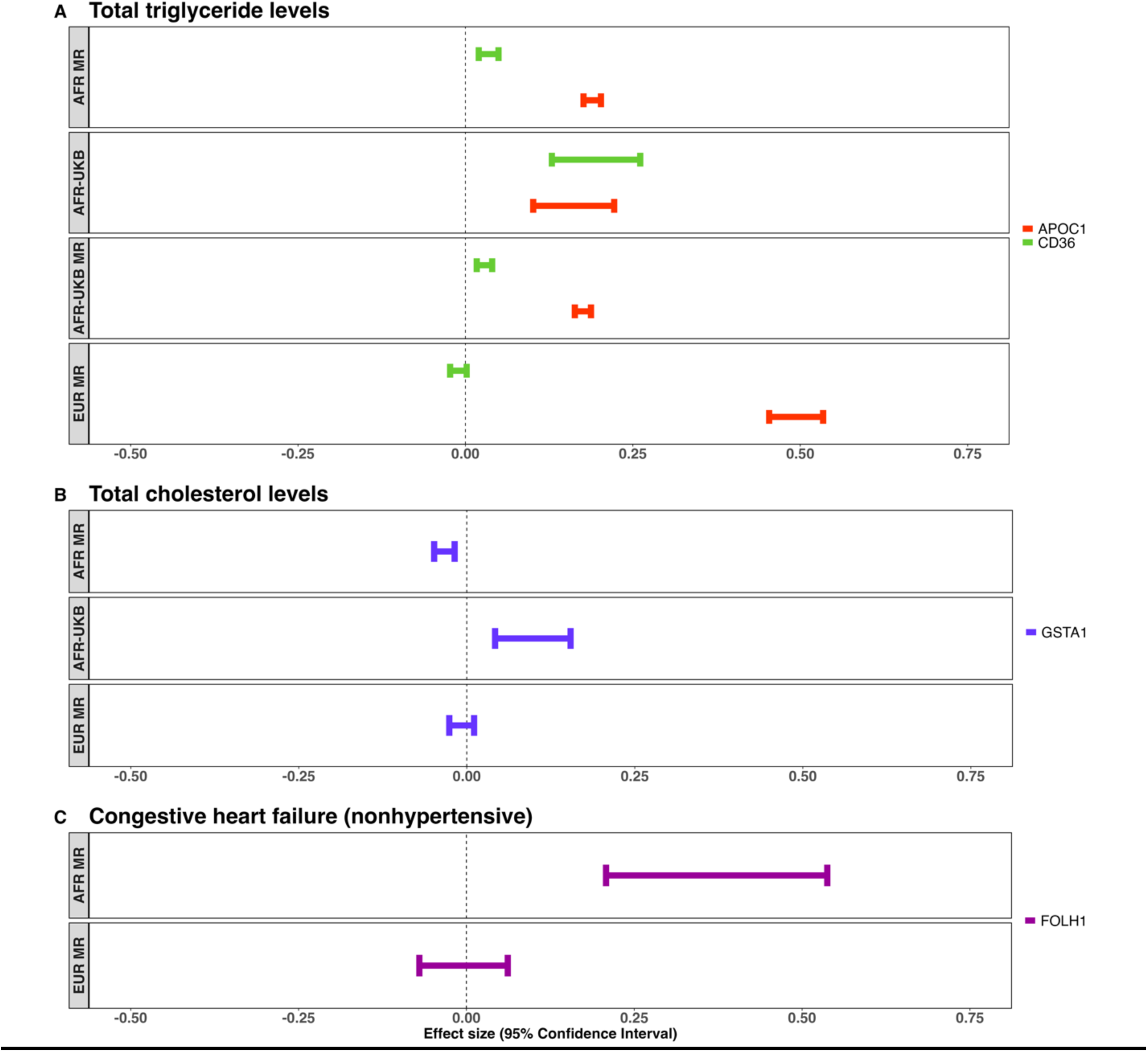
Plot showing differences in absolute effect sizes between African and European populations. Demonstrated in text boxes are differences in absolute effect size for top prioritized protein-outcome associations (**Table 2 and 3**). **Notes**: TG = Triglyceride levels; TC = Total cholesterol; Defibrillator = Cardiac defibrillator in situ; CM = Cardiomyopathy; CHF = Congestive heart failure (non-hypertensive); HDL = High-density lipoproteins; HFrEF = Heart failure with reduced ejection fraction [Systolic or combined heart failure]; HHD = Hypertensive heart disease; PCM = Primary/intrinsic cardiomyopathies; CHF = Congestive heart failure (non-hypertensive).

### Putative natural selection of African ancestry-derived *cis*-pQTLs

We hypothesized that the heterogeneity among protein-outcome associations observed between European- and African-ancestry populations could be the result of evolutionary events. Therefore, to examine whether the *cis*-pQTLs prioritized in the African-ancestry studies showed evidence of natural selection, population branch statistics (PBS) ^75,78,79^ were computed using the 30⨉ whole genome sequencing data from the HGDP. Overall, 95.18% (n=1,540) of the *cis*-pQTLs used in our African MR were present in the HGDP (**Table S19**). Using the HGDP African population, as well as Europeans and East Asians, we computed the PBS of 1,540 *cis*-pQTLs for each population, with the other two populations used as references. We found significant increases of PBS scores using these *cis*-pQTLs in Africans compared to Europeans (p=2.28×10^-13^) and East Asians (p=1.98×10^-27^), exhibiting evidence of natural selection in Africans (**Figure S4A, Table S19**). Moreover, among the 115 significant protein-outcome associations in the African populations (**Table S6**), the average PBS score of 40 unique *cis*-pQTLs demonstrated a 1.6 to 2-fold increase in Africans compared to in Europeans (Wilcoxon test, p=6.4×10^-4^) and East Asians (p=3.1×10^-4^) (**Figure S4B**). Specifically, 19 of 40 of *cis*-pQTLs in the 115 associations had PBS scores greater than 0.1 (mean value of 1,540 *cis*-pQTLs), suggesting that putative natural selection may contribute to the significant MR associations we observed in the African populations.

## Discussion

Drugs developed with the support of human genetics and blood proteomics are more likely to reach approval ^1,5,7,80,81^. This is partially due to the benefits that blood proteomics can provide, which can represent direct, accessible drug targets used to elucidate genetic architecture and disentangle complex bio-mechanisms ^82^. With the recent development of high-plex proteomic platforms, comprehensive mapping of protein-genetic architecture using pQTLs has become feasible. In this study, we leveraged cis-pQTLs in the blood proteome as instruments for MR to uncover causal cardiometabolic disease associations in African-ancestry populations and bring to light modifiable risk factors that could lead to higher disease burden in this population, which is often underserved. Upon conducting MR in African-ancestry populations, and comparing results in Europeans, 52 associations were significant in the African-ancestry population. Among the prioritized associations, four were replicated in the UKB-PPP and 11 exhibited a direct association with plasma protein levels. Ultimately, we found that by MR glycoprotein IIIb (CD36) and apolipoprotein C1 (APOC1) were positively associated with triglyceride levels, glutathione S-transferase alpha 1 (GSTA1) was negatively associated with total cholesterol levels, and folate hydrolase 1 (FOLH1) was positively associated with congestive heart failure (non-hypertensive) in African-ancestry populations.

CD36 was found to be a potential novel therapeutic target because it was positively associated with triglyceride levels demonstrated by MR (b=0.035, se=0.008, p=4.17×10^-6^) and direct associations in the UKB-PPP cohort (b=0.198, CI=0.134-0.263, p=2.43×10^-9^). CD36 was also negatively associated with HDL levels (b=-0.116, se=0.008, p=9.79×10^-54^) by MR, however limited sample sizes may have impacted direct replications in the UKB-PPP (**Table 3**). Interestingly, the effect of CD36 on both triglyceride levels and HDL levels were opposite for European- and African-ancestry populations by MR (**Table S9**), possibly due to the African CD36 pQTL (rs3211938) resulting in a loss-of-function mutation, potentially affecting protein binding. Nonetheless, this variant is an eQTL in multiple tissues (**Table S13**), supporting its role as a pQTL, and was also found to be associated with monocyte surface CD36 and platelet total CD36 receptors in a prior study ^83^. CD36 was also significant by MR for several cardiovascular traits, including cardiomyopathy (b=-0.179, se=0.026, p=2.85×10^-12^), primary/intrinsic cardiomyopathies (b=-0.183, se=0.026, p=2.15×10^-12^), congestive heart failure (non-hypertensive) (b=-0.085, se=0.019, p=5.30×10^-6^), heart failure with reduced EF (systolic or combined heart failure) (b=-0.122, se=0.023, p=1.44 ×10^-7^), hypertensive heart disease (b=-0.121, se=0.024, p=3.56×10^-7^), and implanted cardiac defibrillators (b= −0.193, se=0.040, p=1.82×10^-6^), as well as a phenotypic association among congestive heart failure (non-hypertensive) with plasma CD36 protein levels that did not reach significance. Associations between CD36 and cardiovascular traits have been previously identified, including the relationship between CD36 and cardiac energy metabolism and function. For instance, during heart failure — evidenced by the deletion or deactivation of CD36 — fatty acid metabolism was diminished and resulted in increased glucose metabolism to sustain ATP synthesis and maintain cardiac muscle function ^84-86^. Through dietary supplementation of fatty acids, cardiac dysfunction could be improved, suggesting that CD36-induced fatty acid metabolism is essential for healthy cardiac function and that supplementation represents a useful therapy during heart failure ^84^. Moreover, CD36 deficiency (rs3211938 nonsense mutation) has been previously investigated in Yoruban Africans to discern its association with severe malaria phenotypes ^87,88^, considering the high frequency of this variant in Africans compared to other populations (MAF=0.114; **Table 4**) ^88,89^, as well as the putative selective pressure from endemic exposure to the *Plasmodium falciparum* parasite ^90^. While this variant has not been robustly associated with severe malaria ^87,88,91,92^, the high frequency of rs3211938 in Yorubans and its biological implications remain ambiguous. CD36 deficiency has also been inversely linked to HDL levels in Yorubans in the literature (b=-1.07, se=0.24, p-val=2.36×10^-5^) ^83^ — corroborated by the findings of this study (**Table 3**) — and supporting the decreased susceptibility to metabolic syndromes and cardiovascular diseases ^83,93–96^. A growing body of work also supports that CD36 is an immune-relevant glycoprotein that functions as a receptor for multiple ligands ^83,97,98^. For instance, CD36 has been associated as a fatty acid sensor that controls the uptake and transport of triglyceride-rich lipoproteins from the intestines ^99–101^. Fatty-acid uptake was also shown to be impaired in CD36-deficient disease models, leading to lipid retention ^102^ and increased cardiovascular risks ^99^, suggesting that its role in lipid surveillance normally induces downstream signalling pathways that control dietary lipid intake ^101^. Notably, this was also associated with an increased preference for fatty foods in rodents ^103^ and has been observed in humans carrying different CD36 polymorphisms ^104^. Although not reaching significance in this study, CD36 levels have been found to influence coagulation and thrombosis following COVID-19 infection ^105,106^. That is, CD36-bound COVID-19 envelope proteins — a component associated with viral assembly and budding ^98^ — can recruit mitogen-activated protein kinases (MAPKs) and NF-κB cells to promote platelet activation and augment thrombosis of pulmonary arteries ^105^, remaining present for one year post-infection ^106^. Together, these findings suggest that CD36 acts through several modalities on cardiovascular and vascular risks and may be protective for severe and hospitalized COVID-19 infection by mitigating thrombosis.

**Table 4.** Population branch statistics (PBS) and allele frequency distribution of *cis*-pQTLs of prioritized proteins in African ancestry populations.

| Protein | *cis*-pQTL | PBS | Allele Frequency in HGDP |  |  |
| --- | --- | --- | --- | --- | --- |
|  |  |  | African | East Asian | European |
| AARS | rs3972075 | 0.425 | 0.350 | 0.000 | 0.002 |
| ADH1C | rs1154433 | 0.017 | 0.103 | 0.086 | 0.398 |
| ANGPTL3 | rs35529421 | 0.048 | 0.415 | 0.191 | 0.317 |
| ANXA7 | rs112673115 | 0.156 | 0.159 | 0.000 | 0.008 |
| APOA5 | rs3135506 | 0.000 | 0.065 | 0.001 | 0.065 |
| APOC1 | rs12721054 | 0.160 | 0.150 | 0.000 | 0.000 |
| APOE | rs769455 | 0.022 | 0.025 | 0.000 | 0.000 |
| ASRGL1 | rs1406384 | 0.179 | 0.306 | 0.004 | 0.112 |
| B4GALT1 | rs7019909 | 0.066 | 0.186 | 0.487 | 0.100 |
| BCAM | rs1135062 | 0.088 | 0.439 | 0.110 | 0.317 |
| BCAT2 | rs4802505 | 0.011 | 0.011 | 0.001 | 0.210 |
| C4BPA | rs8942 | 1.130 | 0.891 | 0.168 | 0.186 |
| CBR3 | rs60409141 | 0.034 | 0.502 | 0.388 | 0.354 |
| CCDC126 | rs17148243 | 0.145 | 0.138 | 0.000 | 0.001 |
| CD109 | rs9442950 | 0.201 | 0.184 | 0.000 | 0.000 |
| CD274 | rs7041009 | 0.030 | 0.413 | 0.363 | 0.716 |
| CD36 | rs3211938 | 0.119 | 0.114 | 0.000 | 0.000 |
| CES1 | rs116200666 | 0.071 | 0.129 | 0.012 | 0.039 |
| CROT | rs31659 | 0.014 | 0.128 | 0.264 | 0.101 |
| DDX19A | rs79724048 | 0.464 | 0.377 | 0.000 | 0.004 |
| DLL1 | rs2344744 | 0.109 | 0.412 | 0.021 | 0.550 |
| EFEMP1 | rs4146921 | 0.380 | 0.040 | 0.795 | 0.211 |
| FOLH1 | rs111339910 | 0.062 | 0.062 | 0.000 | 0.000 |
| GAS6 | rs6602911 | 0.270 | 0.210 | 0.540 | 0.596 |
| GATM | rs2668747 | 0.052 | 0.145 | 0.182 | 0.732 |
| GCKR | rs1083864 | 0.778 | 0.102 | 0.820 | 0.589 |
| GLTPD2 | rs11868705 | 0.166 | 0.155 | 0.000 | 0.000 |
| GSTA1 | rs3957356 | 0.037 | 0.328 | 0.155 | 0.421 |
| HP | rs5471 | 0.147 | 0.139 | 0.000 | 0.000 |
| LILRB5 | rs12975366 | 0.034 | 0.156 | 0.114 | 0.448 |
| MANF | rs4687812 | 0.041 | 0.265 | 0.333 | 0.024 |
| PCYOX1 | rs34041544 | 0.074 | 0.074 | 0.000 | 0.000 |
| PLTP | rs6073958 | 0.030 | 0.292 | 0.155 | 0.210 |
| PROC | rs200045749 | 0.012 | 0.015 | 0.000 | 0.000 |
| PROS1 | rs182634126 | 0.025 | 0.027 | 0.000 | 0.000 |
| PTPRJ | rs11039557 | 0.186 | 0.172 | 0.000 | 0.001 |
| SERPINH1 | rs12275382 | 0.116 | 0.114 | 0.000 | 0.002 |
| TIMD4 | rs111873972 | 0.056 | 0.057 | 0.000 | 0.000 |
| UBE2I | rs8063 | 0.483 | 0.658 | 0.041 | 0.280 |
| YWHAH | rs5994430 | 0.154 | 0.394 | 0.150 | 0.144 |

APOC1 was highlighted due to the association with triglyceride levels demonstrated by MR (b=0.189, se=0.007, p=7.98×10^-179^) and direct association with plasma protein levels (b=0.166, CI= 0.102-0.229, p=3.51 ×10^-7^), as well as evidence suggesting its role in cardiovascular diseases and risk factors ^107,108^. For instance, APOC1 functions as an inhibitor of lipoproteins, primarily binding to LDL receptors in the liver and influencing the residence time of APOC1-bound lipoproteins in the plasma during their conversion to LDL^107^. APOC1 has also been associated with hypertriglyceridemia ^109^, possibly due to decreased LDL conversion. Compared to Europeans (MR: b=0.494, se=0.021, p=1.29×10^-127^), it appeared that African-ancestry populations exhibited a lower magnitude of effect, possibly contributing to a lesser risk of atherosclerosis and heart diseases, and potentially representing a weaker therapeutic target. Nonetheless, sample size and power influence the ability to make these direct comparisons.

GSTA1 was negatively associated with cholesterol levels demonstrated by MR (b=-0.033, se=0.008, p=1.79×10^-5^) in African populations, while not significant in Europeans (b= −0.008, se=0.009, p=0.421), however, it was positively associated with plasma protein levels (b=0.098, CI=0.042-0.154, p=6.29×10^-4^) in the observed study in the UKB-PPP among African ancestry populations. GSTA1 is a member of a broad class of enzymes with glutathione transferase activity, catalyzing reactions involved in the hepatic detoxification of drugs, toxins, carcinogens, and products of oxidative stress ^110^. Numerous *GSTA1* polymorphisms have been identified ^110^, exhibiting vast interindividual catalytic activity and pharmacogenetic variation that may influence the metabolism of common drugs such as acetaminophen ^111^. Moreover, a study on the distribution of *GSTA1* promoter haplotypes uncovered SNPs in African populations distinct from other super-population groups, possibly resulting in different GSTA1 metabolic profiles ^112^. Glutathione transferase variants and/or gene expression has been previously associated with conditions such as hypercholesterolemia, hypertriglyceridemia, and low HDL-cholesterol levels, influencing the risk of atherosclerosis and coronary heart disease through factors such as the dietary intake of polyunsaturated fatty acids ^113,114^. The non-concordance observed between MR results and plasma protein levels for GSTA1 in the African ancestry populations remains unclear, which could be the result of cross-platform effects — as no *cis*-pQTLs for GSTA1 in the African populations of UKB-PPP cohort were identified. Altogether, the highly polymorphic nature of *GSTA1* and functional impact of GSTA1 on disease and drug metabolism exemplifies an arduous component of drug development.

FOLH1 was positively associated with congestive heart failure (non-hypertensive) demonstrated by MR (b=0.373, se=0.084, p=9.25×10^-6^) and direct association (b=1.798, CI=1.12-2.48, p=2.20×10^-7^). This result represents a novel association that is not well described in the literature. FOLH1 (also known as prostate-specific membrane antigen) is highly expressed in prostate epithelial tissue and represents a potential biomarker for prostate and bladder cancers ^115,116^.

Additionally, DEAD-box helicase 19A (DDX19A) was associated with both LDL and total cholesterol levels in populations of African ancestry (MR: b=-0.108, se=0.022, p=6.28×10^-7^; b=-0.105, se=0.021, p=6.52×10^-7^, respectively), but not in Europeans (MR: b=-0.012, se=0.011, p=2.60×10^-1^; b=0.025, se=0.031, p=4.12×10^-1^, respectively). Our results also suggest that the prioritized pQTL of DDX19A is potentially under natural selection in African-ancestry populations (rs79724048; PBS=0.464, **Table 4**). DEAD-box helicases are involved in gene expression and are associated with diverse physiological functions^117^. While the specific role of DDX19A is not well described, several DEAD-box helicases have been shown to play a role in cardiovascular diseases through the increased uptake (*e.g*., DDX5) or efflux (*e.g*., DDX17) of lipids, contributing to atherosclerosis or protecting cardiac function, respectively ^118,119^. Moreover, several eQTLs associated with DDX19A have been identified in cardiovascular and vascular tissues (**Table S13**), suggesting it may also contribute to cardiovascular health through an unidentified mechanism. Although not replicated in the UKB-PPP due to the absence of proteomic measurements, MR evidence and the elevated PBS and allele frequency (AF=0.377) in African-ancestry populations may help substantiate DDX19A as a potential candidate for cholesterol level management.

The scope of this investigation is impacted by the limited availability of publicly accessible datasets in diverse populations, as well as the limited power arising from small sample sizes of non-European trait and disease GWASs ^30,120,121^. Likewise, African proteomic GWASs have fewer samples compared to European proteomic GWASs, resulting in fewer proteins with *cis*-pQTLs identified in African-ancestry populations. Moreover, publicly available, non-European GWASs often contain missing variables and are a subset of results or meta-analyses with no ancestry information ^122,123^. This greatly reduced the number of traits and diseases included in our study for African-ancestry populations. The limited number of individuals with African ancestry and proteomic measurements in UKB-PPP could also lead to false negative replication of MR findings. Finally, available reference panels may not be representative of all populations, introducing bias in certain analyses, such as colocalization.

## Conclusions

Overall, multiple lines of evidence were used to triangulate robust proteomic determinants of cardiometabolic diseases and traits with stronger causal influence in African-ancestry populations. We highlighted circulating protein targets that are novel or implicated by previous studies which could represent modifiable risk factors of cardiovascular diseases. This study corroborates evidence toward genetically supported drug targets in African populations and brings forward the importance of diversity for revealing novel therapeutic targets.

## Data Availability

All data used in this study were publicly accessed. A description of all parameters and their origins are described in Supplementary tables 3 and 4.

## Acknowledgments

NA

## Availability of data and materials

All data used in this study were publicly accessed. A description of all parameters and their origins are described in **Supplementary tables 3** and **4**.

## Non-standard Abbreviations and Acronyms

Word: Abbreviation
1KGP: 1000-genomes
AV: Atrioventricular
*cis-*pQTL: *cis-*acting pQTL
DR: Diabetic retinopathy
EAF: Effect allele frequency
EF: Ejection fraction
QTL: eQTL expression
GBMI: Global Biobank Meta-analysis Initiative
HF: Heart failure
HGDP: Human Genome Diversity Project
IPF: Idiopathic pulmonary fibrosis
MR: Mendelian randomization
MAF: Minor allele frequency
NEC: Necrotizing enterocolitis
NOS: Not otherwise specified
PBS: Population branch statistics
QTL: pQTL protein
PAVs: Protein altering variants
QTL: Quantitative trait loci
SOMAmers: SOMAmer reagents
QTL: sQTL splicing
UKB: UK Biobank
VTE: Venous thromboembolism

---

**Supplemental Figure 1:** Forest plots demonstrating MR effect sizes and the direction of effect when comparing the African discovery cohort, the African UKB replication cohort, and the European replication cohort.

**Supplemental Figure 2: a)** FOLH1 protein quantity in congestive heart failure (non-hypertensive) cases and controls from the UKB-PPP (regression estimate and p-value = cases); **b)** APOC1 protein quantity and circulating triglyceride levels linear regression; **c)** CD36 protein quantity and circulating triglyceride levels linear regression; **d)** GSTA1 protein quantity and circulating triglyceride levels linear regression.

**Supplemental Figure 3: a)** BGALT1 protein quantity and circulating LDL levels linear regression); **b)** BCAM protein quantity and circulating LDL levels linear regression; **c)** BCAT1 protein quantity and circulating LDL levels linear regression; **d)** CD36 protein quantity and circulating HDL levels linear regression; **e)** SERPINH1 protein quantity and height levels linear regression.

**Supplemental Figure 4: a)** CD274 protein quantity in unstable angina (intermediate coronary syndrome) cases and controls from the UKB-PPP (regression estimate and p- value = cases); **b)** CD36 protein quantity in congestive heart failure (non- hypertensive)(regression estimate and p-value = cases).

**Supplemental Figure 5: a)** Absolute value of PBS of African (AFR), East Asians (EAS) and Europeans (EUR) of 1,540 cis-pQTLs from African MR. Also showing mean and SD for each population; **b)** Absolute value of PBS of African (AFR), East Asians (EAS) and Europeans (EUR) of 40 cis-pQTLs from 115 prioritized associations (Table S7). Also showing mean and SD for each population.

