## Supplementals2 for "Phenome-wide Mendelian randomization identifying circulating proteins for cardiovascular traits in populations of African ancestry"

**Supplemental material**

(includes supplemental figures, tables, and a description of data tools)

### S1 Figure. MR results of selected significant protein to outcome associations

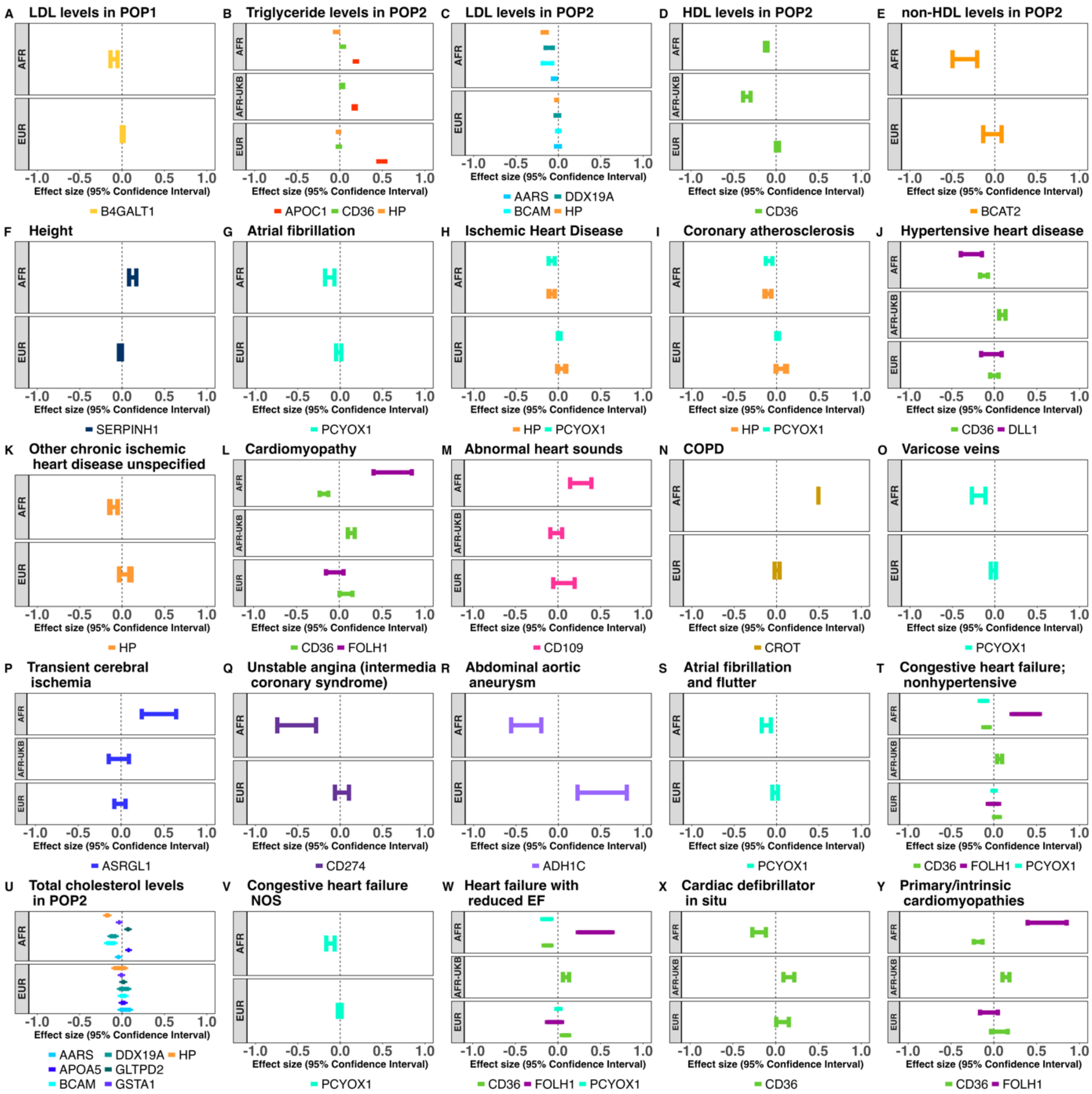


Forest plots demonstrating MR effect sizes and the direction of effect when comparing the African discovery cohort, the African UKB replication cohort, and the European replication cohort.

**S2 Figure. Significant association between protein quantity and phenotypes in the UKB-PPP**

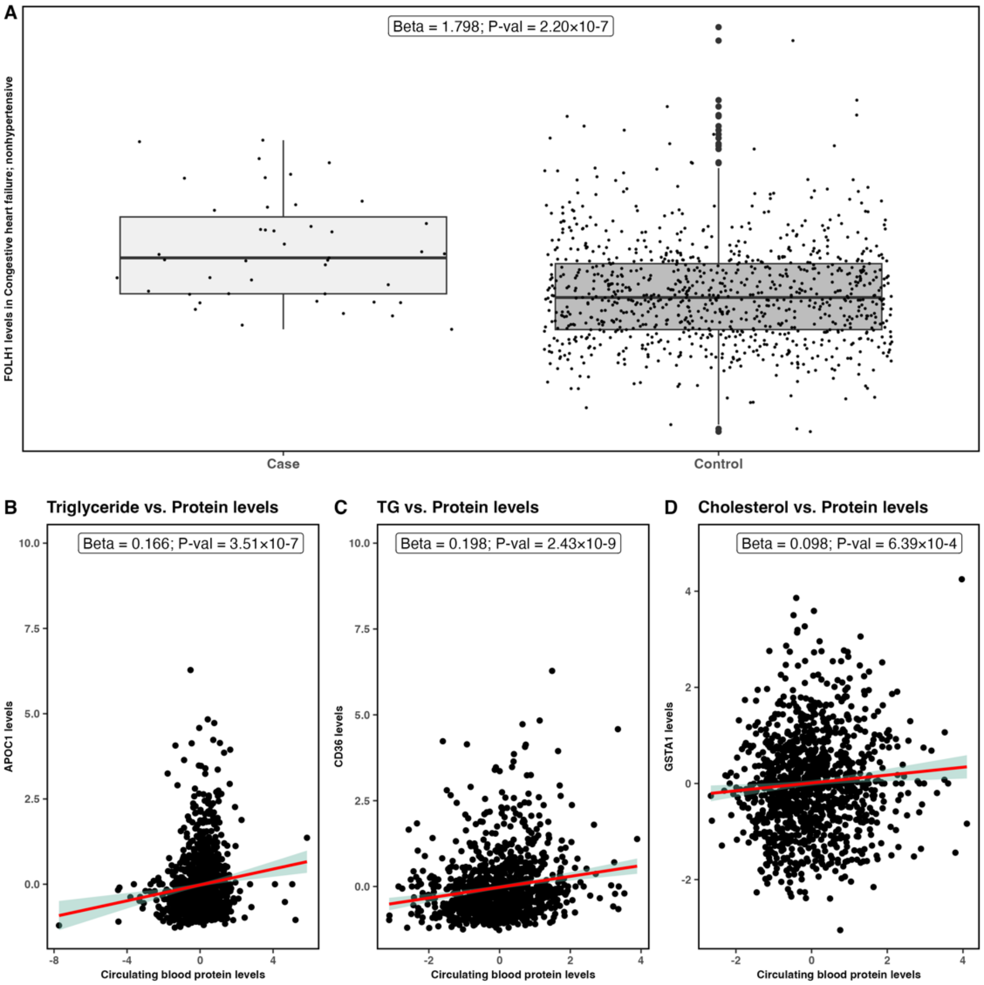


**a)** FOLH1 protein quantity in congestive heart failure (non-hypertensive) cases and controls from the UKB-PPP (regression estimate and p-value = cases); **b)** APOC1 protein quantity and circulating triglyceride levels linear regression; **c)** CD36 protein quantity and circulating triglyceride levels linear regression; **d)** GSTA1 protein quantity and circulating triglyceride levels linear regression.

**S3 Figure. Other association between protein quantity and continuous phenotypes in the UKB-PPP**

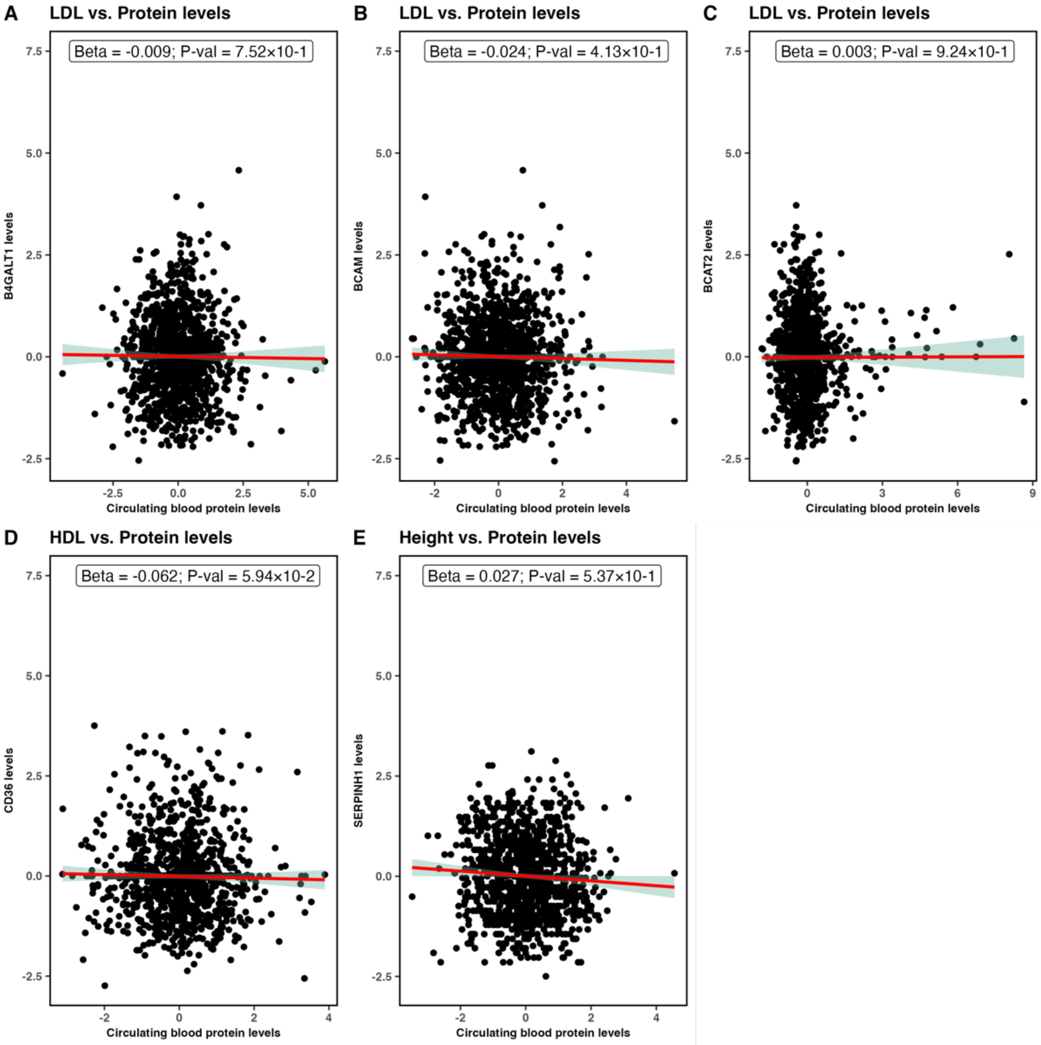


**a)** BGALT1 protein quantity and circulating LDL levels linear regression); **b)** BCAM protein quantity and circulating LDL levels linear regression; **c)** BCAT1 protein quantity and circulating LDL levels linear regression; **d)** CD36 protein quantity and circulating HDL levels linear regression; **e)** SERPINH1 protein quantity and height levels linear regression.

**S4 Figure. Other association between protein quantity and binary phenotypes in the UKB-PPP**

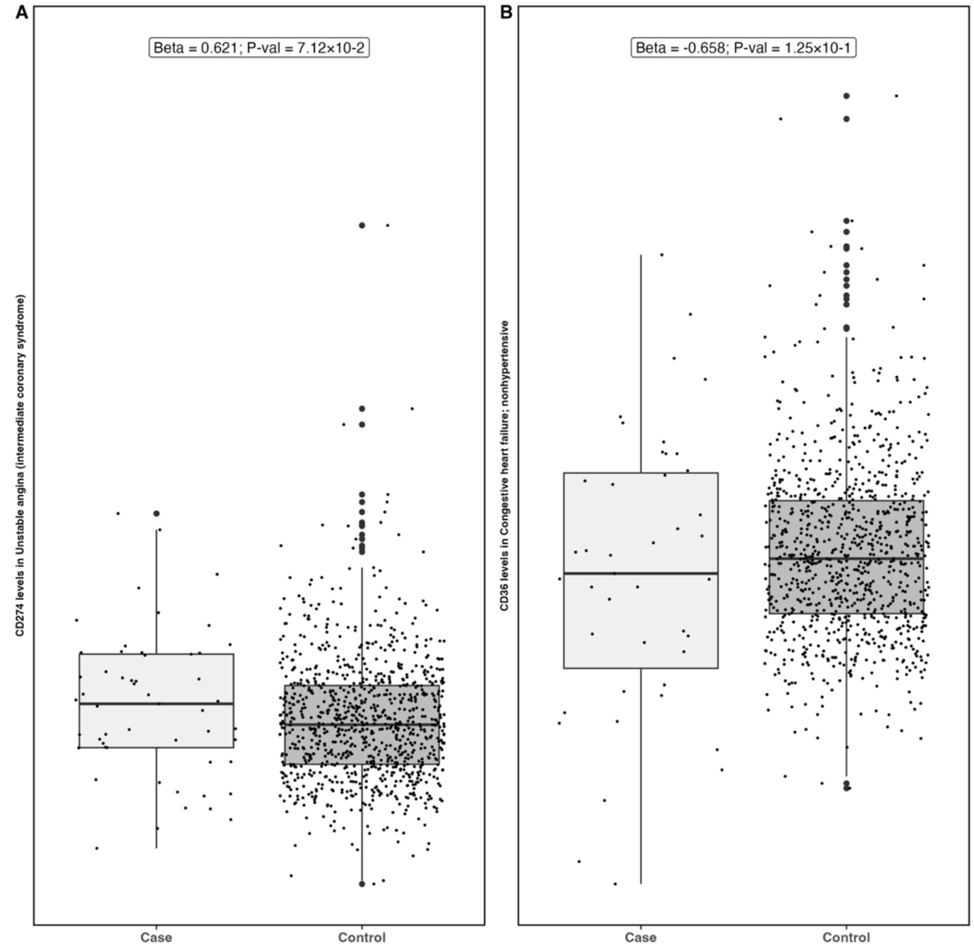


**a)** CD274 protein quantity in unstable angina (intermediate coronary syndrome) cases and controls from the UKB-PPP (regression estimate and p-value = cases); **b)** CD36 protein quantity in congestive heart failure (non-hypertensive)(regression estimate and p-value = cases).

**S5 Figure. PBS of cis-pQTLs from African-ancestry MR**

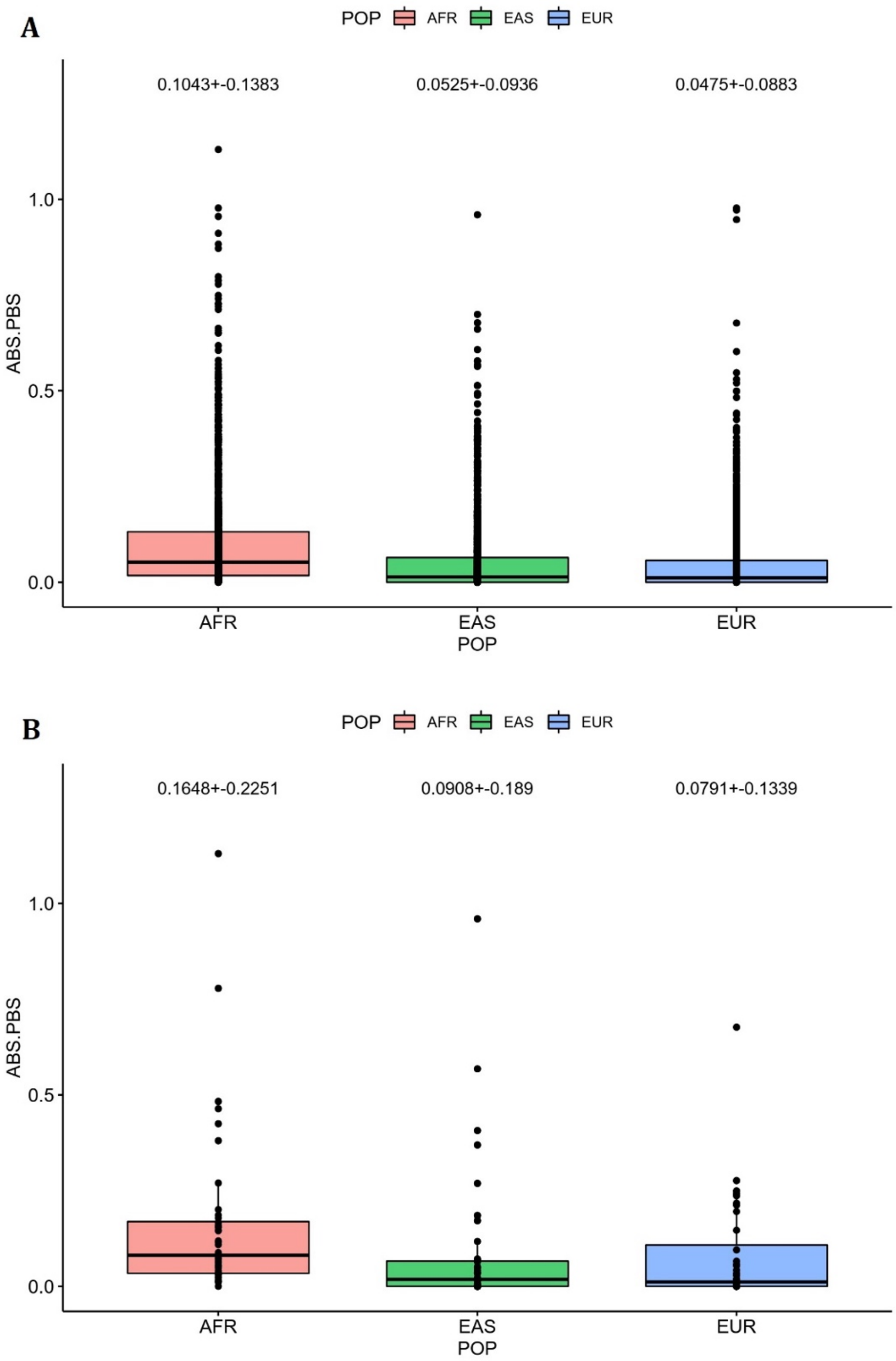


**a)** Absolute value of PBS of African (AFR), East Asians (EAS) and Europeans (EUR) of 1,540 cis-pQTLs from African MR. Also showing mean and SD for each population; **b)** Absolute value of PBS of African (AFR), East Asians (EAS) and Europeans (EUR) of 40 cis-pQTLs from 115 prioritized associations (Table S7). Also showing mean and SD for each population.

### S1 Appendix. Outcome GWAS Summary of African Ancestry Populations

| Trait | n | | | Population description | GWAS catalog ascension number | PMID |
|---|---|---|---|---|---|---|
| | Total | Cases | Controls | | | |
| **Asthma** | 4182 | 2638 | 1544 | POP2 | GCST90131434 | 36089080 |
| **BMI** | 27610 | 27610 | NA | POP2 | GCST008124 | 29273807 |
| **COPD** | 14804 | 1263 | 13541 | POP4 | NA | 36777996 |
| **Very severe respiratory confirmed COVID-19 vs. population** | 13196 | 628 | 12568 | POP4 | NA | 34237774 |
| **Hospitalized COVID-19 vs. population** | 125814 | 2589 | 123225 | POP4 | NA | 34237774 |
| **COVID-19 vs. population** | 137954 | 8814 | 129140 | POP4 | NA | 34237774 |
| **DR vs. no DR** | 1852 | 911 | 941 | POP3 | NA | 30487263 |
| **Gout** | 46099 | 2202 | 43897 | POP4 | NA | 36777996 |
| **HF** | 39685 | 1742 | 37943 | POP4 | NA | 36777996 |
| **Height** | 41389 | 41389 | NA | POP3 | GCST90013466 | 33713608 |
| **IPF** | 17592 | 362 | 17230 | POP4 | NA | 36777996 |
| **LDL** | 24515 | 24515 | NA | POP1 | GCST90101745 | 35546142 |
| **LDL** | 99432 | 99432 | NA | POP2 | GCST90239656 | 34887591 |
| **HDL** | 24616 | 24616 | NA | POP1 | GCST90101746 | 35546142 |
| **HDL** | 99432 | 99432 | NA | POP2 | GCST90239650 | 34887591 |
| **Total cholesterol** | 24612 | 24612 | NA | POP1 | GCST90101747 | 35546142 |
| **Total cholesterol** | 99432 | 99432 | NA | POP2 | GCST90239674 | 34887591 |
| **Triglyceride levels** | 24600 | 24600 | NA | POP1 | GCST90101748 | 35546142 |
| **Triglyceride levels** | 99432 | 99432 | NA | POP2 | GCST90239662 | 34887591 |
| **Non-HDL cholesterol levels** | 99432 | 99432 | NA | POP2 | GCST90239668 | 34887591 |
| **PTSD** | 9223 | 2479 | 6744 | POP3 | NA | 28439101 |
| **POAG** | 4441 | 2320 | 2121 | POP2 | GCST009245 | 31688885 |
| **Stroke** | 23991 | 3961 | 20030 | POP2 | GCST90104549 | 36180795 |
| **Cardioembolic stroke** | 20271 | 241 | 20030 | POP2 | GCST90104551 | 36180795 |
| **Large artery stroke** | 20243 | 213 | 20030 | POP2 | GCST90104552 | 36180795 |
| **Small vesicle stroke** | 20453 | 423 | 20030 | POP2 | GCST90104553 | 36180795 |
| **Ischemic stroke** | 20924 | 894 | 20030 | POP2 | GCST90104550 | 36180795 |
| **Smoking initiation (ever vs. never)** | 10558 | 4261 | 6297 | POP1, POP4 | GCST90091238 | 36335192 |
| **Smoking cessation (current vs. former)** | 4257 | 2471 | 1786 | POP1, POP4 | GCST90091239 | 36335192 |

| | | | | | | |
|---|---|---|---|---|---|---|
| **T2D** | 4347 | 2633 | 1714 | POP1 | GCST008114 | 31049640 |
| **VTE** | 32508 | 1466 | 31042 | POP4 | NA | 36777996 |
| **Abdominal aortic aneurysm** | 120603 | 1773 | 118830 | POP3 | NA | 26441289 |
| **Abnormal electrocardiogram** | 107978 | 10831 | 97147 | POP3 | NA | 26441289 |
| **Abnormal function study of cardiovascular system** | 117968 | 1289 | 116679 | POP3 | NA | 26441289 |
| **Abnormal heart sounds** | 115967 | 3258 | 112709 | POP3 | NA | 26441289 |
| **Acute pulmonary heart disease** | 120829 | 3605 | 117224 | POP3 | NA | 26441289 |
| **Acute but ill-defined cerebrovascular disease** | 120049 | 4568 | 115481 | POP3 | NA | 26441289 |
| **Angina pectoris** | 113926 | 7533 | 106393 | POP3 | NA | 26441289 |
| **Aortic aneurysm** | 120250 | 2825 | 117425 | POP3 | NA | 26441289 |
| **Arrhythmia (cardiac) NOS** | 111419 | 8648 | 102771 | POP3 | NA | 26441289 |
| **Arterial embolism and thrombosis** | 120649 | 958 | 119691 | POP3 | NA | 26441289 |
| **Atherosclerosis** | 116040 | 5248 | 110792 | POP3 | NA | 26441289 |
| **Atherosclerosis of native arteries of the extremities with intermittent claudication** | 119068 | 2794 | 116274 | POP3 | NA | 26441289 |
| **Atherosclerosis of native arteries of the extremities with ulceration or gangrene** | 121279 | 1050 | 120229 | POP3 | NA | 26441289 |
| **Atherosclerosis of the extremities** | 118254 | 4282 | 113972 | POP3 | NA | 26441289 |
| **Atrial fibrillation** | 118847 | 8873 | 109974 | POP3 | NA | 26441289 |
| **Atrial fibrillation and flutter** | 118682 | 9482 | 109200 | POP3 | NA | 26441289 |
| **Atrial flutter** | 120231 | 3470 | 116761 | POP3 | NA | 26441289 |
| **Atrioventricular block** | 119057 | 2893 | 116164 | POP3 | NA | 26441289 |
| **Atrioventricular block complete** | 121500 | 856 | 120644 | POP3 | NA | 26441289 |
| **Blood vessel replaced** | 120991 | 694 | 120297 | POP3 | NA | 26441289 |
| **Bundle branch block** | 119151 | 2051 | 117100 | POP3 | NA | 26441289 |
| **Cardiac arrest** | 120967 | 582 | 120385 | POP3 | NA | 26441289 |
| **Cardiac arrest and ventricular** | 120680 | 828 | 119852 | POP3 | NA | 26441289 |
| **Cardiac conduction disorders** | 107083 | 17288 | 89795 | POP3 | NA | 26441289 |

| | | | | | | |
|---|---|---|---|---|---|---|
| **Cardiac defibrillator *in situ*** | 121503 | 2629 | 118874 | POP3 | NA | 26441289 |
| **Cardiac dysrhythmias** | 104362 | 33210 | 71152 | POP3 | NA | 26441289 |
| **Cardiac pacemaker *in situ*** | 121037 | 2348 | 118689 | POP3 | NA | 26441289 |
| **Cardiac pacemaker/device *in situ*** | 121094 | 4153 | 116941 | POP3 | NA | 26441289 |
| **Cardiomegaly** | 115275 | 3619 | 111656 | POP3 | NA | 26441289 |
| **Cardiomyopathy** | 118560 | 7053 | 111507 | POP3 | NA | 26441289 |
| **Carditis** | 119409 | 2074 | 117335 | POP3 | NA | 26441289 |
| **Cerebral artery occlusion with cerebral infarction** | 119017 | 7521 | 111496 | POP3 | NA | 26441289 |
| **Cerebral ischemia** | 116795 | 8197 | 108598 | POP3 | NA | 26441289 |
| **Cerebrovascular disease** | 114381 | 16858 | 97523 | POP3 | NA | 26441289 |
| **Chronic pulmonary heart disease** | 118672 | 3527 | 115145 | POP3 | NA | 26441289 |
| **Chronic venous hypertension** | 121505 | 703 | 120802 | POP3 | NA | 26441289 |
| **Chronic venous insufficiency** | 117614 | 5227 | 112387 | POP3 | NA | 26441289 |
| **Circulatory disease NEC** | 116323 | 3033 | 113290 | POP3 | NA | 26441289 |
| **Congestive heart failure NOS** | 116880 | 11137 | 105743 | POP3 | NA | 26441289 |
| **Congestive heart failure; non-hypertensive** | 116379 | 15405 | 100974 | POP3 | NA | 26441289 |
| **Coronary atherosclerosis** | 114151 | 20733 | 93418 | POP3 | NA | 26441289 |
| **Deep vein thrombosis** | 119901 | 4478 | 115423 | POP3 | NA | 26441289 |
| **Elevated blood pressure reading without diagnosis of hypertension** | 109712 | 8192 | 101520 | POP3 | NA | 26441289 |
| **Encounter for long-term (current) use of anticoagulants antithrombotic aspirin** | 118328 | 1880 | 116448 | POP3 | NA | 26441289 |
| **Encounter for long-term (current) use of aspirin** | 118593 | 1557 | 117036 | POP3 | NA | 26441289 |
| **Endocarditis** | 120734 | 772 | 119962 | POP3 | NA | 26441289 |
| **Essential hypertension** | 116845 | 93857 | 22988 | POP3 | NA | 26441289 |
| **First degree AV block** | 119719 | 1415 | 118304 | POP3 | NA | 26441289 |
| **Heart failure NOS** | 119845 | 1451 | 118394 | POP3 | NA | 26441289 |

| | | | | | | |
|---|---|---|---|---|---|---|
| **Heart failure with preserved EF [Diastolic heart failure]** | 118420 | 5379 | 113041 | POP3 | NA | 26441289 |
| **Heart failure with reduced EF [Systolic or combined heart failure]** | 118736 | 9104 | 109632 | POP3 | NA | 26441289 |
| **Heart transplant/surgery** | 121064 | 518 | 120546 | POP3 | NA | 26441289 |
| **Heart valve disorders** | 116021 | 5334 | 110687 | POP3 | NA | 26441289 |
| **Heart valve replaced** | 121587 | 931 | 120656 | POP3 | NA | 26441289 |
| **Hemorrhoids** | 102646 | 17547 | 85099 | POP3 | NA | 26441289 |
| **Hypertension** | 116935 | 94292 | 22643 | POP3 | NA | 26441289 |
| **Hypertensive chronic kidney disease** | 115875 | 14127 | 101748 | POP3 | NA | 26441289 |
| **Hypertensive heart and/or renal disease** | 112649 | 22462 | 90187 | POP3 | NA | 26441289 |
| **Hypertensive heart disease** | 114864 | 8634 | 106230 | POP3 | NA | 26441289 |
| **Hypotension** | 111927 | 9298 | 102629 | POP3 | NA | 26441289 |
| **Hypotension NOS** | 114148 | 5728 | 108420 | POP3 | NA | 26441289 |
| **Iatrogenic hypotension** | 119955 | 584 | 119371 | POP3 | NA | 26441289 |
| **Ill-defined descriptions and complications of heart disease** | 114186 | 3053 | 111133 | POP3 | NA | 26441289 |
| **Intracerebral hemorrhage** | 121477 | 557 | 120920 | POP3 | NA | 26441289 |
| **Intracranial hemorrhage** | 121142 | 1349 | 119793 | POP3 | NA | 26441289 |
| **Ischemic Heart Disease** | 110452 | 26859 | 83593 | POP3 | NA | 26441289 |
| **Late effects of cerebrovascular disease** | 118994 | 5530 | 113464 | POP3 | NA | 26441289 |
| **Left bundle branch block** | 120502 | 879 | 119623 | POP3 | NA | 26441289 |
| **Myocardial infarction** | 117239 | 7835 | 109404 | POP3 | NA | 26441289 |
| **Non-infectious disorders of lymphatic channels** | 120674 | 1249 | 119425 | POP3 | NA | 26441289 |
| **Nonrheumatic aortic valve disorders** | 119594 | 2397 | 117197 | POP3 | NA | 26441289 |
| **Nonrheumatic mitral valve disorders** | 118056 | 2682 | 115374 | POP3 | NA | 26441289 |
| **Nonspecific abnormal findings on radiologic imaging of heart and coronary circulation** | 117284 | 1215 | 116069 | POP3 | NA | 26441289 |
| **Nonspecific chest pain** | 103924 | 42507 | 61417 | POP3 | NA | 26441289 |
| **Occlusion and stenosis of precerebral arteries** | 117082 | 3388 | 113694 | POP3 | NA | 26441289 |

| | | | | | | |
|---|---|---|---|---|---|---|
| **Occlusion of cerebral arteries** | 118880 | 7610 | 111270 | POP3 | NA | 26441289 |
| **Orthostatic hypotension** | 116944 | 2837 | 114107 | POP3 | NA | 26441289 |
| **Other aneurysm** | 119640 | 3536 | 116104 | POP3 | NA | 26441289 |
| **Other chronic ischemic heart disease unspecified** | 116620 | 11327 | 105293 | POP3 | NA | 26441289 |
| **Other disorders of arteries and arterioles** | 119522 | 1243 | 118279 | POP3 | NA | 26441289 |
| **Other disorders of circulatory system** | 115215 | 4127 | 111088 | POP3 | NA | 26441289 |
| **Other forms of chronic heart disease** | 117307 | 4726 | 112581 | POP3 | NA | 26441289 |
| **Other hypertensive complications** | 117304 | 3465 | 113839 | POP3 | NA | 26441289 |
| **Other specified cardiac dysrhythmias** | 110488 | 9339 | 101149 | POP3 | NA | 26441289 |
| **Other specified peripheral vascular disease** | 120600 | 769 | 119831 | POP3 | NA | 26441289 |
| **Other venous embolism and thrombosis** | 118315 | 7556 | 110759 | POP3 | NA | 26441289 |
| **Palpitations** | 115735 | 5606 | 110129 | POP3 | NA | 26441289 |
| **Paroxysmal supraventricular tachycardia** | 120134 | 1843 | 118291 | POP3 | NA | 26441289 |
| **Paroxysmal tachycardia unspecified** | 118810 | 3882 | 114928 | POP3 | NA | 26441289 |
| **Paroxysmal ventricular tachycardia** | 120056 | 2003 | 118053 | POP3 | NA | 26441289 |
| **Pericarditis** | 120533 | 1246 | 119287 | POP3 | NA | 26441289 |
| **Peripheral vascular disease** | 115762 | 9993 | 105769 | POP3 | NA | 26441289 |
| **Peripheral vascular disease unspecified** | 116126 | 9303 | 106823 | POP3 | NA | 26441289 |
| **Phlebitis and thrombophlebitis** | 119664 | 1130 | 118534 | POP3 | NA | 26441289 |
| **Precordial pain** | 119573 | 609 | 118964 | POP3 | NA | 26441289 |
| **Premature beats** | 117656 | 2106 | 115550 | POP3 | NA | 26441289 |
| **Primary pulmonary hypertension** | 120969 | 608 | 120361 | POP3 | NA | 26441289 |
| **Primary/intrinsic cardiomyopathies** | 118885 | 6758 | 112127 | POP3 | NA | 26441289 |
| **Pulmonary embolism and infarction acute** | 120880 | 3545 | 117335 | POP3 | NA | 26441289 |
| **Pulmonary heart disease** | 118408 | 7085 | 111323 | POP3 | NA | 26441289 |
| **Rheumatic disease of the heart valves** | 118726 | 1230 | 117496 | POP3 | NA | 26441289 |

| | | | | | | |
|---|---|---|---|---|---|---|
| **Right bundle branch block** | 120438 | 1024 | 119414 | POP3 | NA | 26441289 |
| **Second degree AV block** | 121368 | 536 | 120832 | POP3 | NA | 26441289 |
| **Secondary/extrinsic cardiomyopathies** | 120668 | 783 | 119885 | POP3 | NA | 26441289 |
| **Sinoatrial node dysfunction (Bradycardia)** | 121007 | 695 | 120312 | POP3 | NA | 26441289 |
| **Subdural hemorrhage** | 121698 | 542 | 121156 | POP3 | NA | 26441289 |
| **Supraventricular premature beats** | 120013 | 556 | 119457 | POP3 | NA | 26441289 |
| **Symptoms involving cardiovascular system** | 118005 | 1256 | 116749 | POP3 | NA | 26441289 |
| **Tachycardia NOS** | 113676 | 5094 | 108582 | POP3 | NA | 26441289 |
| **Transient cerebral ischemia** | 117310 | 7606 | 109704 | POP3 | NA | 26441289 |
| **Unstable angina (intermediate coronary syndrome)** | 117568 | 3328 | 114240 | POP3 | NA | 26441289 |
| **Varicose veins** | 118376 | 3597 | 114779 | POP3 | NA | 26441289 |
| **Varicose veins of lower extremity** | 119567 | 2757 | 116810 | POP3 | NA | 26441289 |
| **Varicose veins of lower extremity symptomatic** | 120405 | 1622 | 118783 | POP3 | NA | 26441289 |

**Definitions**: Atrioventricular (AV); Body mass index (BMI); Chronic obstructive pulmonary disease (COPD); Diabetic retinopathy (DR); Ejection fraction (EF); Heart failure (HF); High-density lipoprotein (HDL); Idiopathic pulmonary fibrosis (IPF); Low-density lipoprotein (LDL); Necrotizing enterocolitis (NEC); not otherwise specified (NOS);Post-traumatic stress disorder (PTSD); Primary open-angle glaucoma (POAG); Type II diabetes (T2D); Venous thromboembolism (VTE); Africa Wits-INDEPTH Partnership for Genomic Studies (AWI-Gen); Africa America Diabetes Mellitus study (AADM); The COVID-19 host genetics initiative (COVID-hg); Durban Diabetes Study (DDS); Durban Diabetes Case Control study (DCC); Genetic Investigation of Anthropometric Traits (GIANT); Global Biobank Meta-analysis Initiative (GBMI); Global Lipids Genetics Consortium (GLGC); The Psychiatric Genomics Consortium-Posttraumatic Stress Disorder group (PGC-PTSD); The Psychiatric Genomics Consortium (PGC); The Uganda Genome Resource (UGR); Individuals of African ancestry in UK Biobank (UKB-AFR); POP1 = Sub-Saharan African; POP2 = African American or Afro-Caribbean or African-unspecified; POP3 = African-American; POP4 = African-unspecified.

**S2 Appendix. Corresponding European MR results of prioritized associations in African-ancestry populations (Table 1)**

| Number of *cis*-pQTLs | Protein/ Gene | Outcome | Effect allele | Other allele | Effect allele Frequency | MR Beta | MR SE | MR p-value |
|---|---|---|---|---|---|---|---|---|
| 1 | AARS | LDL | A | T | 0.9732 | -0.006 | 0.011 | $6.30\times10^{-1}$ |
| | | Total cholesterol | A | T | 0.9732 | 0.039 | 0.034 | $2.58\times10^{-1}$ |
| 1 | ADH1C | Abdominal aortic aneurysm | T | C | 0.033 | 0.516 | 0.148 | $4.96\times10^{-4}$ |
| 2 | APOA5 | Total cholesterol | C<br>A | G<br>G | 0.0619<br>0.9386 | 0.013 | 0.015 | $3.82\times10^{-1}$ |
| 2 | APOC1 | Triglyceride levels | C<br>T | G<br>C | 0.4627<br>0.7655 | 0.494 | 0.021 | $1.29\times10^{-127}$ |
| 2 | ASRGL1 | Transient cerebral ischemia | A<br>C | T<br>G | 0.7242<br>0.8824 | -0.014 | 0.033 | $6.83\times10^{-1}$ |
| 1 | B4GALT1 | LDL | T | C | 0.1048 | 0.008 | 0.006 | $1.61\times10^{-1}$ |
| 1 | BCAM | LDL | A | G | 0.9896 | -0.001 | 0.007 | $8.86\times10^{-1}$ |
| | | Total cholesterol | A | G | 0.9896 | 0.012 | 0.020 | $5.56\times10^{-1}$ |
| 2 | BCAT2 | non-HDL cholesterol | C<br>A | G<br>G | 0.0183<br>0.2186 | -0.028 | 0.055 | $6.14\times10^{-1}$ |
| 5 | CD109 | Abnormal heart sounds | C<br>T<br>C<br>T<br>A | G<br>G<br>G<br>G<br>G | 0.5105<br>0.0436<br>0.0164<br>0.9804<br>0.0191 | 0.070 | 0.064 | $3.53\times10^{-1}$ |
| 2 | CD274 | Unstable angina (intermediate coronary syndrome) | T<br>A | C<br>C | 0.2515<br>0.09 | 0.020 | 0.042 | $6.29\times10^{-1}$ |
| 2 | CD36 | Triglyceride levels | | | | -0.010 | 0.006 | $9.63\times10^{-2}$ |
| | | Cardiac defibrillator *in situ* | | | | 0.081 | 0.038 | $3.10\times10^{-2}$ |
| | | Cardiomyopathy | T | C | 0.4069 | 0.078 | 0.039 | $4.18\times10^{-2}$ |
| | | Congestive heart failure; non-hypertensive | | | | 0.037 | 0.017 | $3.20\times10^{-2}$ |
| | | HDL | | | | 0.016 | 0.009 | $8.08\times10^{-2}$ |
| | | Heart failure with reduced EF [Systolic or combined heart failure] | A | G | 0.9315 | 0.089 | 0.022 | $5.33\times10^{-5}$ |
| | | Hypertensive heart disease | | | | 0.001 | 0.025 | $9.74\times10^{-1}$ |
| | | Primary/intrinsic cardiomyopathies | | | | 0.065 | 0.050 | $1.89\times10^{-1}$ |
| 1 | CROT | COPD | A | G | 0.1076 | 0.006 | 0.015 | $6.86\times10^{-1}$ |
| 1 | DDX19A | LDL | A | G | 0.9873 | -0.012 | 0.011 | $2.60\times10^{-1}$ |
| | | Total cholesterol | | | | 0.025 | 0.031 | $4.12\times10^{-1}$ |
| 1 | DLL1 | Hypertensive heart disease | A | C | 0.578 | -0.034 | 0.062 | $5.82\times10^{-1}$ |
| 1 | FOLH1 | Congestive heart failure; non-hypertensive | T | C | 0.9487 | -0.004 | 0.034 | $8.99\times10^{-1}$ |

| Number of *cis*-pQTLs | Protein/ Gene | Outcome | Effect allele | Other allele | Effect allele Frequency | MR Beta | MR SE | MR p-value |
|---|---|---|---|---|---|---|---|---|
| | | Cardiomyopathy | | | | -0.053 | 0.052 | $3.15\times10^{-1}$ |
| | | Heart failure with reduced EF [Systolic or combined heart failure] | | | | -0.040 | 0.045 | $3.72\times10^{-1}$ |
| | | Primary/intrinsic cardiomyopathies | | | | -0.057 | 0.053 | $2.87\times10^{-1}$ |
| 3 | GLTPD2 | Total cholesterol | A<br>T<br>T | G<br>G<br>C | 0.3521<br>0.143<br>0.0166 | 0.014 | 0.011 | $1.93\times10^{-1}$ |
| 1 | GSTA1 | Total cholesterol | A | C | 0.6421 | -0.008 | 0.009 | $4.21\times10^{-1}$ |
| 12 | HP* | Coronary atherosclerosis | T | C | 0.1982 | 0.060 | 0.032 | $1.55\times10^{-1}$ |
| | | Coronary atherosclerosis | T | C | 0.1982 | 0.044 | 0.029 | $1.83\times10^{-1}$ |
| | | Ischemic Heart Disease | A | G | 0.1266 | 0.044 | 0.025 | $1.70\times10^{-1}$ |
| | | Ischemic Heart Disease | T | G | 0.8754 | 0.032 | 0.024 | $2.37\times10^{-1}$ |
| | | LDL | A | G | 0.052 | -0.021 | 0.002 | $1.11\times10^{-29}$ |
| | | LDL | A | G | 0.052 | -0.022 | 0.002 | $2.00\times10^{-28}$ |
| | | Other chronic ischemic heart disease unspecified | A | G | 0.9565 | 0.041 | 0.036 | $3.44\times10^{-1}$ |
| | | Other chronic ischemic heart disease unspecified | T | C | 0.0162 | 0.025 | 0.031 | $4.50\times10^{-1}$ |
| | | Total cholesterol | A | G | 0.9493 | -0.039 | 0.005 | $4.49\times10^{-13}$ |
| | | Total cholesterol | A | G | 0.9493 | -0.042 | 0.005 | $1.69\times10^{-14}$ |
| | | Total cholesterol | A | G | 0.0308 | -0.027 | 0.038 | $5.28\times10^{-1}$ |
| | | Triglyceride levels<br>Triglyceride levels | T | C | 0.0184 | -0.015<br>-0.013 | 0.004<br>0.004 | $3.37\times10^{-5}$<br>$1.26\times10^{-4}$ |
| 2 | PCYOX1 | Varicose veins | T<br>T | C<br>C | 0.1499<br>0.9339 | -0.017 | 0.016 | $2.78\times10^{-1}$ |
| 1 | SERPINH1 | Height | A | T | 0.4205 | -0.019 | 0.009 | $2.91\times10^{-2}$ |

**Notes**: * = multiple SOMAmers; **Definitions**: Chronic obstructive pulmonary disease (COPD); High-density lipoprotein (HDL); Ejection fraction (EF); Low-density lipoprotein (LDL).

### **Data packages, tools, software, webtools, and URLs**:

| Tool/Program/Package | | Reference/URL |
|---|---|---|
| **R v4.1.2** | **TwoSampleMR v0.5.8** | Hemani G., *et al*. 2018. The MR-Base Collaboration. The MR-Base platform supports systematic causal inference across the human phenome. eLife. 7:e34408. doi: 10.7554/eLife.34408. https://mrcieu.github.io/TwoSampleMR/ |
| **R v4.1.2** | **Coloc v5.2.1** | Giambartolomei C., *et al*. 2014. Bayesian Test for Colocalisation between Pairs of Genetic Association Studies Using Summary Statistics. PLOS Genetics. https://doi.org/10.1371/journal.pgen.1004383. https://chr1swallace.github.io/coloc/ |
| **Snappy v1.0** | | Forgetta V. (2022). richardslab/snappy: Snappy: A flexible SNP proxy finder (v1.1). Zenodo. https://doi.org/10.5281/zenodo.7328428 https://gitlab.com/richards-lab/vince.forgetta/snappy |
| **Python v3.11.4** | **scikit-allel v1.3.8** | Miles A., *et al*. 2024. cggh/scikit-allel: v1.3.8 (v1.3.8). Zenodo. https://doi.org/10.5281/zenodo.10876220 https://github.com/cggh/scikit-allel |
| **UCSC** | **Liftover** | Kent W.J., *et al*. 2002. The human genome browser at UCSC. Genome Res. 12(6):996-1006. https://genome.ucsc.edu/cgi-bin/hgLiftOver http://genome.ucsc.edu |
| **UCSC** | **hg38 chain files** | |
| **UCSC** | **hg19 chain files** | |
| **PLINK v1.9** | | Purcell S. *et al.* PLINK: a toolset for whole-genome association and population-based linkage analysis. Journal of Human Genetics. http://pngu.mgh.harvard.edu/purcell/plink/ |
| **METAL** | | Willer C.J., *et al*. 2016. METAL: fast and efficient meta-analysis of genomewide association scans. Bioinformatics. 1;26(17):2190-1. doi: 10.1093/bioinformatics/btq340. Epub 2010 Jul 8. https://csg.sph.umich.edu/abecasis/Metal/index.html |
| **METAL-Random v0.1.0** | | Hemani G. 2022. explodecomputer/random-metal: Adding random effects model (v0.1.0). Zenodo. https://doi.org/10.5281/zenodo.6974696 https://github.com/explodecomputer/random-metal/tree/v0.1.0 |
| **HaploReg v4.2** | | Ward LD, Kellis M.2011. HaploReg: a resource for exploring chromatin states, conservation, and regulatory motif alterations within sets of genetically linked variants. Nucleic Acids Res. 40(Database issue):D930-4. https://pubs.broadinstitute.org/mammals/haploreg/haploreg.php |
| **GTex** | | The data used for the analyses described in this manuscript were obtained from the GTEx Portal on 01/20/2024 and/or dbGaP accession number phs000424.vN.pN |
| **Phenoscanner v2** | | Kamat M.A., *et al*. 2019. PhenoScanner V2: an expanded tool for searching human genotype-phenotype associations. Bioinformatics. 1;35(22):4851-4853. doi: 10.1093/bioinformatics/btz469. http://www.phenoscanner.medschl.cam.ac.uk/ |
| **OMIM** | | Online Mendelian Inheritance in Man, OMIM®. McKusick-Nathans Institute of Genetic Medicine, Johns Hopkins University (Baltimore, MD). May 2024. World Wide Web URL: https://omim.org/ |
| **Genome Reference Panels** | **Human Genetics Diversity Panel (HGDP)** | Fairley S., *et al.* 2020. The International Genome Sample Resource (IGSR) collection of open human genomic variation resources. Nucleic Acids Res. 8;48(D1):D941-D947. doi: 10.1093/nar/gkz836. https://www.internationalgenome.org/data-portal/data-collection/hgdp |
| **Genome Reference Panels** | **gnomADv3.1** | Chen S. *et al*. 2024. A genomic mutational constraint map using variation in 76,156 human genomes Nature 625, 92–100. |

| | Tool/Program/Package | Reference/URL |
|---|---|---|
| | | https://doi.org/10.1038/s41586-023-06045-0 https://gnomad.broadinstitute.org/ |
| **GWAS Resources** | **GWAS catalog** | Sollis E, *et al*. 2022. The NHGRI-EBI GWAS Catalog: knowledgebase and deposition resource. Nucleic Acids Res. gkac1010. doi: 10.1093/nar/gkac1010. Epub ahead of print. PMID: 36350656. https://www.ebi.ac.uk/gwas/home |
|  | **Global biobank meta-analysis initiative (GBMI)** | Zhou W, *et al*. 20220. Global Biobank Meta-analysis Initiative: Powering genetic discovery across human disease. Cell Genom. 12;2(10):100192. doi: 10.1016/j.xgen.2022.100192. https://www.globalbiobankmeta.org/ |
| | **COVID-19 host genetics imitative (HGI) round 7** | The COVID-19 Host Genetics Initiative, a global initiative to elucidate the role of host genetic factors in susceptibility and severity of the SARS-CoV-2 virus pandemic. European Journal of Human Genetics. 2020;28(6):715–718. doi:10.1038/s41431-020-0636-6<br>COVID-19 Host Genetics Initiative. A second update on mapping the human genetic architecture of COVID-19. Nature. 2023;621(7977):E7–E26. doi:10.1038/s41586-023-06355-3<br>https://www.covid19hg.org/ |
| **Biobanks** | **UK Biobank Pharma Proteomics Project (UKB-PPP)** | UK Biobank Pharma Proteomics Project (UKB-PPP) was accessed on DATE from https://registry.opendata.aws/ukbppp. Sun, B.B., *et al*. 2023. Plasma proteomic associations with genetics and health in the UK Biobank. Nature. 622, 329–338. https://doi.org/10.1038/s41586-023-06592-6 |
